# Effects of BNT162b2 mRNA vaccine on Covid-19 infection and hospitalisation among older people: matched case control study for England

**DOI:** 10.1101/2021.04.19.21255461

**Authors:** Thomas Mason, Matt Whitston, Jack Hodgson, Ruth E Watkinson, Yiu-Shing Lau, Omnia Abdulrazeg, Matt Sutton

**Affiliations:** Performance Analysis Team, NHS England & NHS Improvement, Quarry House, Leeds, UK; Health Organisation, Policy & Economics, School of Health Sciences, University of Manchester, UK

**Keywords:** COVID-19, Vaccines, Infections, Observational study

## Abstract

**Background:** The BNT162b2 mRNA vaccine has been shown to be effective at preventing serious Covid-19 events in clinical trials. There is less evidence on effectiveness in real-world settings, especially for older people. The rapid roll-out of the NHS vaccination programme in England based on age thresholds offers an opportunity to make unbiased comparisons of outcomes between vaccinated and unvaccinated populations.

**Methods and Findings:** We matched older (aged 80-83 years) vaccine recipients with younger (aged 76-79 years) persons not yet eligible to receive the vaccine on gender, area of residence, area deprivation, health status, living arrangements, acute illness, and history of seasonal flu vaccination. We also adjusted for the over-representation of Covid-19 positive individuals in the control population because eligibility for vaccination required no Covid-19 symptoms in the previous two weeks. The study population included 170,226 individuals between the ages of 80 and 83 years from community settings outside care homes who received one dose of BNT162b2 mRNA between the 15^th^ and 20^th^ December 2020 and were scheduled a second dose 21 days later.

We found emergency hospital admissions were 51.0% (95% confidence interval: 19.9% to 69.5%) lower and positive Covid-19 tests were 55.2% (40.8% to 66.8%) lower for vaccinated individuals compared to matched controls 21 to 27 days after first vaccination. Emergency admissions were 75.6% (52.8% to 87.6%) lower and positive Covid-19 tests were 70.1% (55.1% to 80.1%) lower 35 to 41 days after first vaccination when 79% of participants had received a second dose within 26 days of their first dose.

**Conclusions:** Receipt of the BNT162b2 mRNA vaccine is effective at reducing Covid-19 hospitalisations and infections. The nationwide vaccination of older adults in England with the BNT162b2 mRNA vaccine reduced the burden of Covid-19.

## Introduction

Several vaccines against severe acute respiratory syndrome coronavirus 2 (SARS-CoV-2) infection and the resulting coronavirus disease 2019 (Covid-19) have been demonstrated to be safe and highly effective in phase 3 randomised clinical trials, with efficacy estimates for prevention of symptomatic disease ranging from 62% to 95%.^1–4^ However, it is also important to examine their effectiveness when deployed in mass vaccination campaigns across diverse populations, where trial exclusion criteria do not apply and where deviations from dosing and handling protocols may occur.

Early evidence from an observational study of mass vaccination using the BNT162b2 mRNA Covid-19 vaccine in Israel estimated real-world effectiveness consistent with reported trial efficacy.^3,5^ This indirectly provided evidence that vaccine effectiveness was maintained against the more transmissible B.1.1.7. variant,^5,6^ which was widespread in the population during the study period. Vaccine effectiveness estimates were consistent across age groups, though were slightly lower amongst people with multiple coexisting health conditions.^5^ Similarly, estimates from Scotland^7^ and England^8^ provide further early evidence of effectiveness. However, such observational, non-randomised studies may be biased by systematic differences between intervention and control groups and between those receiving the intervention at different points in time. The remarkable speed of Covid-19 vaccination rollouts^5,7,8^ and specific prioritisation of vulnerable groups^9^ heighten the risk of these biases, as acknowledged in existing studies.^5,7,8^

We exploit age-based eligibility phasing in the early stages of the nationwide NHS population vaccination programme in England to estimate the real-world effectiveness of the BNT162b2 mRNA vaccine. We match vaccinated persons aged 80 to 83 years to younger persons who did not become eligible for the vaccine until three weeks later and compare their rates of Covid-19 infection and hospitalisation over the 45 days following the date of their first dose. It is particularly important to assess real-world vaccine effectiveness in this older population group, because severe Covid-19 is strongly age-associated^10^ and adaptive immune responses decline with age.^11^

## Methods

### Data

MW and JH obtained population-wide person-level data for England, including vaccination details (date, type and dose), SARS-CoV-2 tests (date, result), age, gender, area of residence, use of hospital services and dates of death. For data sources, linkage methods and access, see Appendix 1. The study population was defined at mid-November 2020. Data were extracted on 9^th^ February 2021 and include complete records to 3^rd^ February 2021.

### Study design

Concurrent with the vaccination programme, COVID-19 (B.1.1.7. variant) incidence in England varied widely, as the B.1.1.7. variant spread through the population^6^ and a national lockdown was implemented.

The first phase of the NHS England vaccination programme targeted: (1) front-line health and social care workers, (2) older care home residents and their carers, and (3) people aged 80 years and over^9^. Differences in risks of exposure and outcome within the first two groups are not effectively measured in administrative datasets. We focused on 171,931 individuals aged 80-83 years not living in care homes that received their first dose between 15^th^ and 20^th^ December 2020, of whom 78.8% received a second dose within 26 days.

We compared vaccinated cases to people aged 72-79 years who became eligible for vaccination later. The speed of vaccination rollout meant many of these received a first vaccine dose during the follow-up period (see Figure 1). We matched vaccinated cases to suitable controls separately for each day of the follow-up period, excluding as controls individuals vaccinated more than two weeks before the day for which outcomes were being compared.

**Figure 1.**
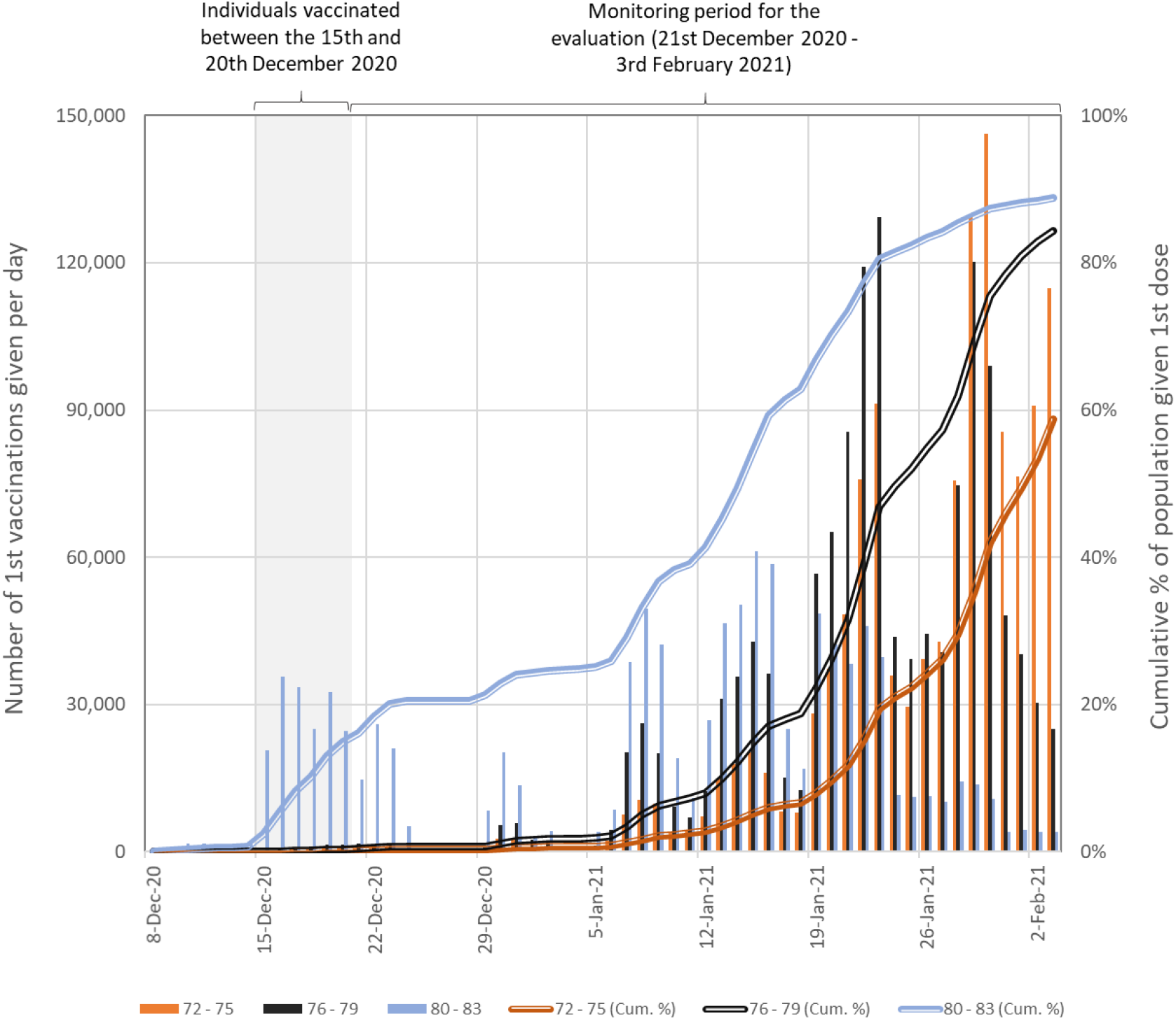
Numbers of people in England who received their first COVID-19 vaccination dose between the 8th December 2020 and the 3rd February 2021 by age group. The cumulative totals are relative to estimates of eligible population based on extracts from the National Health Application and Infrastructure Services (NHAIS) system as of the 15th November 2020. Prior to the 4^th^ January 2021 all individuals received the BNT162b2 mRNA vaccine, after which individuals were vaccinated with either the BNT162b2 mRNA or ChAdOx1 adenovirus vector vaccines.

As individuals should not have had a COVID-19 infection in the two weeks prior to vaccination, the control group contains a progressively higher proportion of people who test positive for COVID-19. This selection process biases the rate of positive tests in the control group upwards and would artificially inflate estimated vaccine effectiveness. To correct for this bias, we sequentially adjusted event rates in the intervention and control groups so that they remained consistent in the first 11 days of follow-up, regardless of the date by which the control group could not (yet) have been vaccinated. Details of the adjustment method are provided in Appendix 2.

To explore adjustment robustness and the sensitivity of the results to the age group used for the controls, we compared the older age group to two different younger age groups. First, we matched vaccinated individuals aged 80-81 to controls aged 76-77, and vaccinated individuals aged 82-83 to controls aged 78-79. Second, we matched vaccinated individuals aged 80-81 to controls aged 72-73, and vaccinated individuals aged 82-83 to controls aged 74-75. The younger control group is less similar in age but unexposed to the vaccine for longer and therefore less prone to selection bias.

We examined rates per 100,000 people for three outcomes: SARS-CoV-2 infection and Covid-19 related hospital attendances and hospitalisations. Infection was recorded by specimen date of SARS-CoV-2 positive polymerase-chain-reaction (PCR) test results from health care facilities (called Pillar 1 testing) or community testing (Pillar 2). Covid-19 related A&E attendances were measured using diagnosis information from emergency department records combined with linked positive Covid-19 test results from 14 days before to 6 days after the attendance (see Appendix 3). Covid-19 related hospital admissions were measured using admitted patient care spell records available on discharge, limited to lengths of stay of 42 days or less. The required information is available for 95% of all emergency department attendances and 93% of all emergency admissions.

### Matching and statistical analysis

Isolating the impact of vaccination requires a study design that accounts for temporal changes in infection rates, for which we used 1:1 exact matching^12^ to account for several factors associated with exposure and outcomes: gender; area of residence;^13^ small area deprivation;^14^ ethnic group; health status; living arrangements; seasonal influenza vaccine history since April 2020; and emergency hospital stays in the previous two months. We excluded 1,705 (1.0%) individuals with prior Covid-19 history to avoid likely pre-existing immunity.^15,16^ We excluded individuals from the control group if they were living in care homes or were not alive on the 15^th^ of December 2020. We dropped matched pairs where either individual was in hospital on the vaccination date or the pair lived at the same property (see Figure 2).

**Figure 2.**
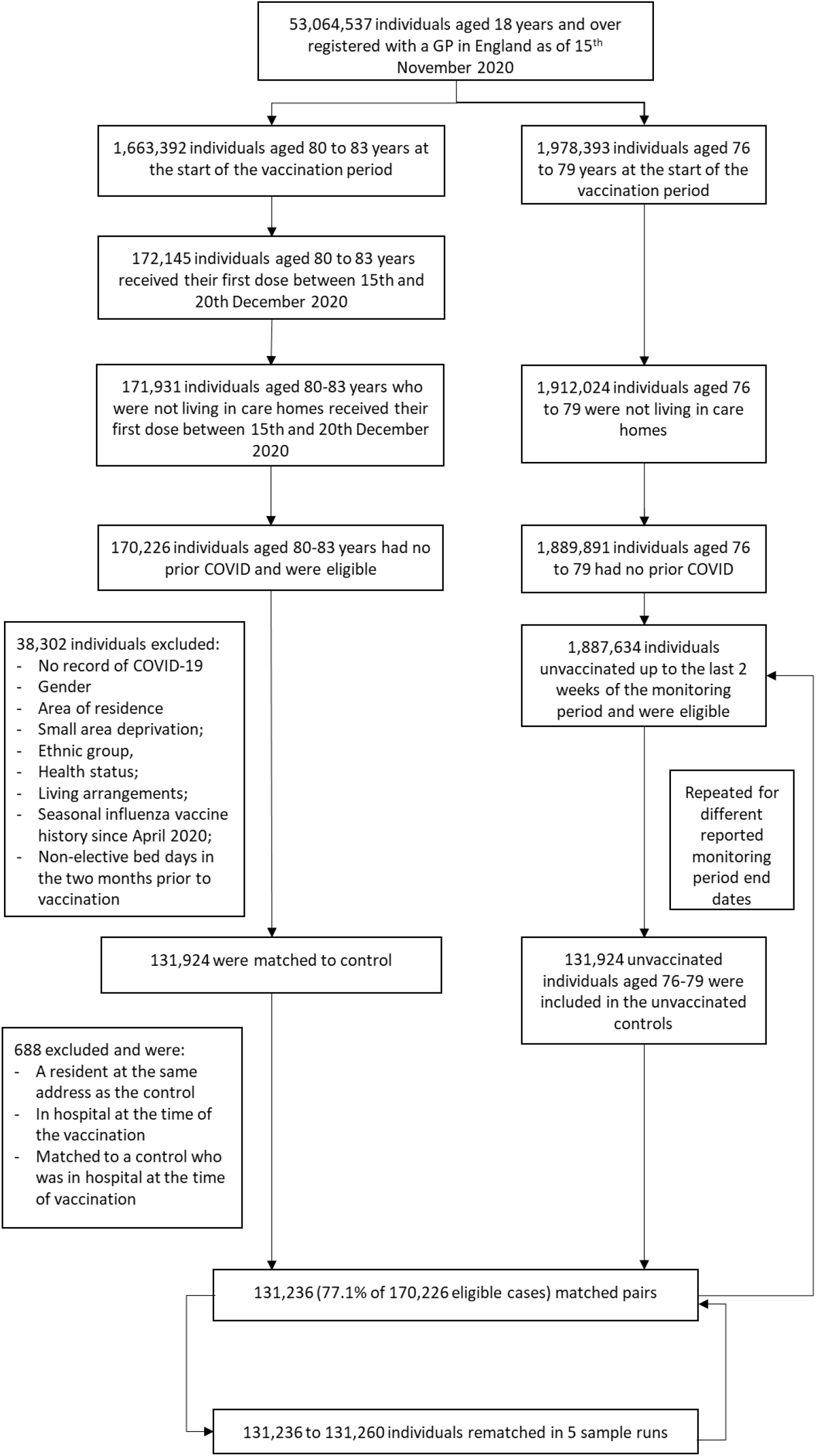
Flow diagram of the study population with eligibility criteria, exclusions and matching methodology.

We matched vaccinated individuals to unique controls without replacement. We assigned a control randomly where multiple matches were available for a vaccinated individual. We repeated the matching process five times with different random number seeds to create five matched populations. We bootstrapped 100 samples with replacement from each of these five matched populations and obtained confidence intervals using percentile values from the 500 samples.

We used the STROBE cohort checklist when writing our report.^17^

## Results

### Study population

Of the total 1,685,530 individuals aged 80-83, 170,226 were not residents of care homes, had no prior history of Covid-19, and received a first dose of the BNT162b2 mRNA vaccine between 15^th^-20^th^ December 2020, so were eligible for inclusion in the study (Figure 2). Of these, we exact-matched 131,236 (77.1%) to control individuals aged 76-79 who were not yet eligible for vaccination (Figures 1&2). The requirement for an exact match generated a matched study population with lower proportions of individuals who were frail or clinically extremely vulnerable, from minority ethnic groups, or from socially-deprived areas compared to the full study population (Table 1 & Appendix 4).

**Table 1.** Demographic and clinical characteristics of vaccinated persons and their unvaccinated controls based on the matched cohort at baseline (day 11 after vaccination).

| Matching criteria | Categories | Total number of 80 to 83 year olds | Total number of 80 to 83 year olds vaccinated | Of which tested positive for COVID-19 | Number with a pairwise control match | % with a pairwise match | % given a second dose by day 45 |
| --- | --- | --- | --- | --- | --- | --- | --- |
| Gender | Female | 869,792 | 89,930 | 153 | 69,819 | 77.6% | 80.6% |
|  | Male | 793,599 | 80,295 | 162 | 61,417 | 76.5% | 80.6% |
|  | Unknown | <5 | <5 | - | - | 0.0% |  |
| Ethnicity | White | 1,375,449 | 157,896 | 273 | 127,605 | 80.8% | 80.5% |
|  | Black | 27,390 | 1,567 | <5 | 270 | 17.2% | 78.8% |
|  | Pakistani and Bangladeshi | 16,466 | 1,141 | 15 | 212 | 18.6% | 86.4% |
|  | Other BAME | 55,180 | 5,922 | 23 | 1,803 | 30.4% | 85.6% |
|  | Unknown | 188,907 | 3,700 | <5 | 1,346 | 36.4% | 80.3% |
| IMD Quintile | 1 (most deprived) | 272,565 | 23,292 | 42 | 17,135 | 73.6% | 86.6% |
|  | 2 | 304,696 | 28,556 | 64 | 20,119 | 70.5% | 81.7% |
|  | 3 | 347,849 | 35,785 | 71 | 26,927 | 75.2% | 80.4% |
|  | 4 | 368,308 | 40,339 | 75 | 31,648 | 78.5% | 79.5% |
|  | 5 (least deprived) | 368,876 | 42,213 | 62 | 35,407 | 83.9% | 78.3% |
|  | Not recorded | 1,098 | 41 | <5 | - | 0.0% |  |
| Living arrangements | Living with children | 127,582 | 5,297 | 23 | 1,224 | 23.1% | 81.7% |
|  | Living alone | 555,343 | 49,876 | 74 | 33,730 | 67.6% | 81.5% |
|  | All other living arrangements | 980,467 | 115,053 | 218 | 96,282 | 83.7% | 80.3% |
| Health Risk | Frail and/or clinically extremely vulnerable | 317,750 | 42,856 | 128 | 28,903 | 67.4% | 81.1% |
|  | Clinically vulnerable | 393,749 | 57,742 | 95 | 43,515 | 75.4% | 80.1% |
|  | Healthy/other | 951,893 | 69,628 | 92 | 58,818 | 84.5% | 80.7% |
| Flu vaccine | Received seasonal flu vaccine FY2020/21 | 948,840 | 150,300 | 263 | 118,746 | 79.0% | 80.3% |
|  | Did not receive seasonal flu vaccine FY2020/22 | 714,552 | 19,926 | 52 | 12,490 | 62.7% | 83.1% |
| Recent acute illness | High | 14,697 | 809 | 7 | 22 | 2.7% | 52.4% |
|  | Medium | 8,701 | 521 | <5 | 6 | 1.2% | 100.0% |
|  | Low | 1,639,994 | 168,896 | 305 | 131,208 | 77.7% | 80.6% |
| Region | East Midlands | 149,390 | 8,830 | 13 | 7,211 | 81.7% | 63.1% |
|  | East of England | 193,482 | 21,805 | 53 | 17,308 | 79.4% | 78.5% |
|  | London | 173,844 | 18,923 | 87 | 10,901 | 57.6% | 85.8% |
|  | North East | 89,680 | 9,300 | 8 | 7,314 | 78.6% | 89.8% |
|  | North West | 231,339 | 22,898 | 31 | 18,312 | 80.0% | 87.7% |
|  | South East | 279,419 | 29,011 | 64 | 22,674 | 78.2% | 81.6% |
|  | South West | 192,927 | 21,349 | 19 | 17,319 | 81.1% | 81.6% |
|  | West Midlands | 182,345 | 22,168 | 19 | 17,678 | 79.7% | 68.9% |
|  | Yorkshire and The Humber | 169,606 | 15,882 | 20 | 12,519 | 78.8% | 87.5% |
|  | Unknown | 1,360 | 60 | <5 | - | 0.0% |  |
| **Total** |  | **1,663,392** | **170,226** | **315** | **131,236** | **77.1%** | **80.6%** |

### Vaccine effectiveness

Across 45 days of follow-up, there were an average of 13.7 documented SARS-CoV-2 infections per day per 100,000 vaccinated individuals, compared to 23.2 per 100,000 unvaccinated controls. Over the same period, a daily average of 5.0 individuals per 100,000 attended A&E with Covid-19 and 5.3 per 100,000 were hospitalised with Covid-19 amongst the vaccinated cohort, compared to 9.6 per 100,000 (attended) and 9.4 per 100,000 (hospitalised) amongst unvaccinated controls.

For the unvaccinated comparison group, event rates increased in the first two weeks of follow-up, with documented infections reaching a maximum at day 20, and emergency hospital attendances and admissions peaking between days 23 and 26 (Figure 3). These profiles reflect the shape of the COVID-19 pandemic in England where prevalence peaked around the 1^st^ January 2021,^18^ and hospitalisations peaked in the second week of January 2021.^19^ For the vaccinated group, documented infections peaked earlier (day 14) and hospitalisations peaked between days 23 and 26. We found similar results when we matched vaccinated individuals to unvaccinated individuals aged 72-75 years, and when the comparison group was restricted to individuals who remained unvaccinated throughout the follow-up period (Appendix 5).

**Figure 3.**
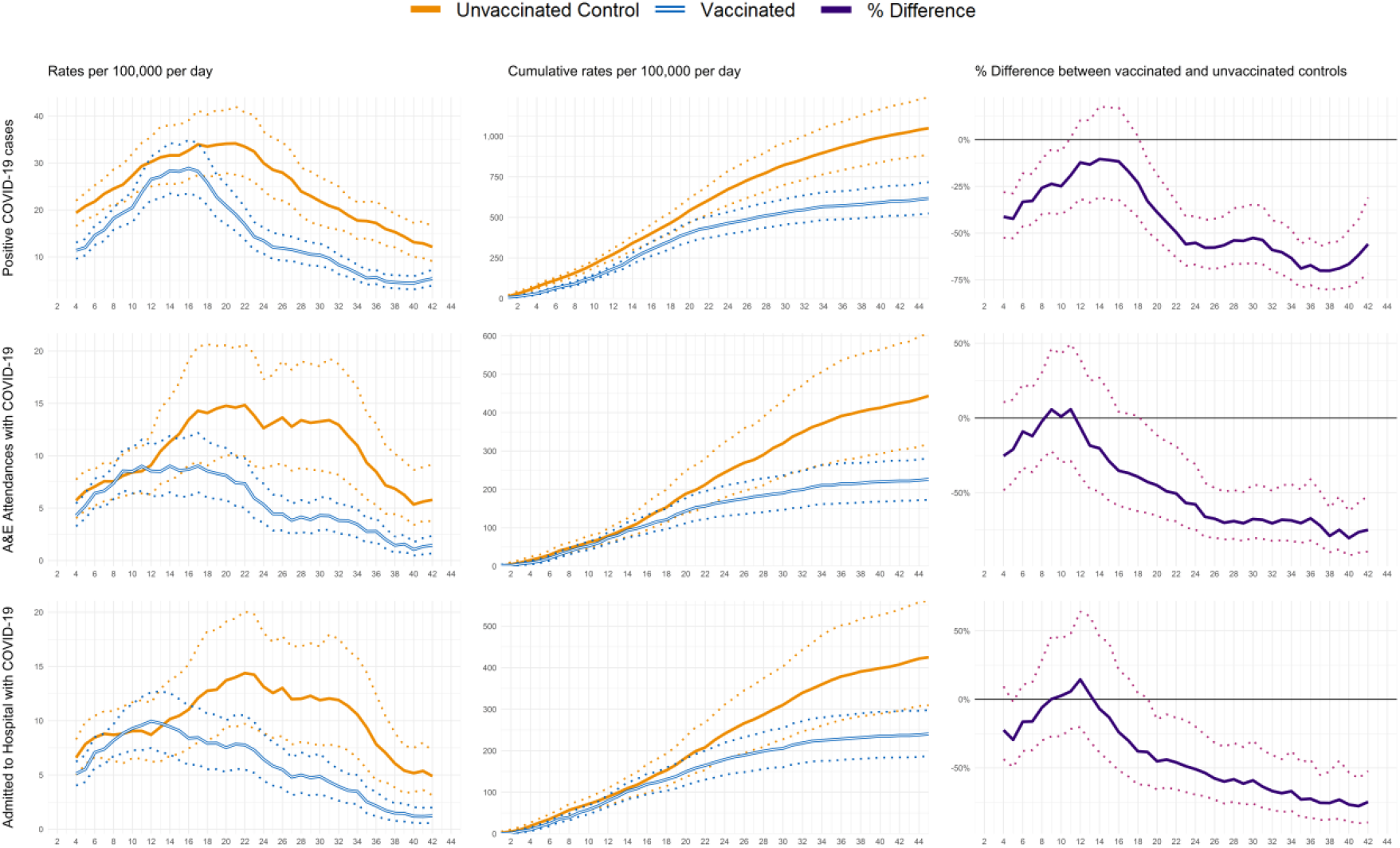
Profiles of positive COVID-19 infections and emergency hospital attendances and admissions by days since first dose of vaccination. The data represent people aged between 80 to 83 years who received their first dose of the BNT162b2 mRNA Covid-19 vaccine between the 15^th^ and 20^th^ December 2020 with comparison to their matched controls. 95% confidence intervals are displayed as dashed lines.

Table 2 shows vaccine effectiveness, defined as percentage difference between vaccinated and unvaccinated groups, for each outcome across four time-periods. Effectiveness increased over the follow-up period for all three outcomes. Estimated effectiveness at 21-27 days was 55.2% (95%CI 40.8%-66.8%) for documented infection, 57.8% (30.8%-74.5%) for emergency hospital attendances, and 50.1% (19.9%-69.5%) for admissions. By day 35-41, estimated effectiveness was 70.1% (55.1%-80.1%) for documented infection, 78.9% (60.0%-89.9%) for emergency hospital attendances, and 75.6% (52.8%-87.6%) for hospitalisations.

**Table 2.**
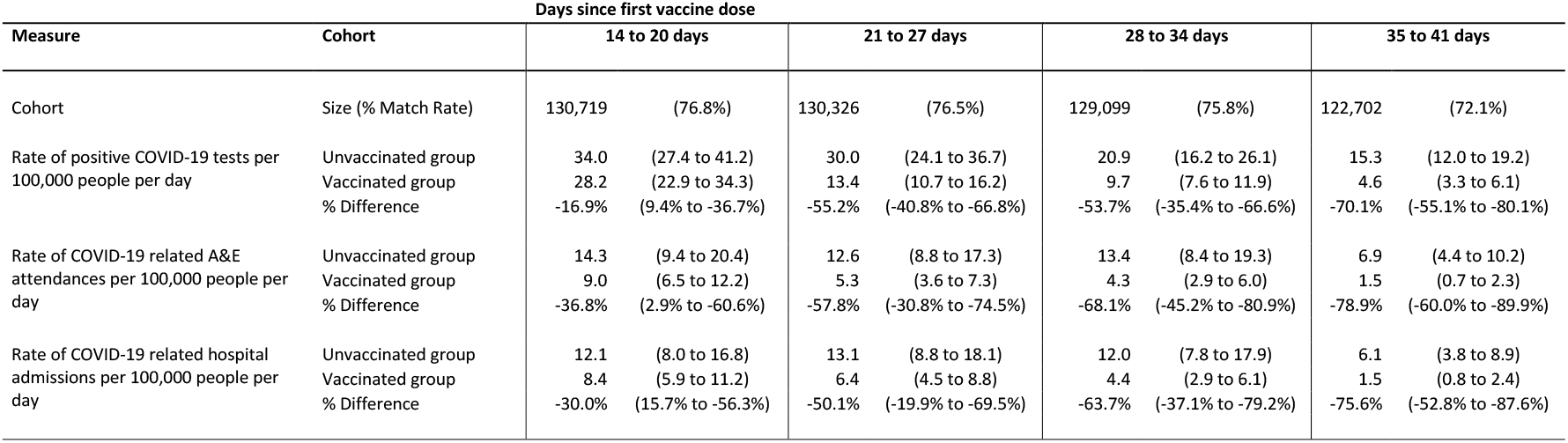
Estimates of the effectiveness of the BNT162b2 mRNA Covid-19 vaccine by days since vaccination.

## Discussion

### Statement of principal findings

We compared rates of SARS-CoV-2 positive tests and Covid-19 hospitalisations in the 45-day period after 171,931 individuals aged 80 to 83 years in England received a first dose of BNT162b2 mRNA Covid-19 vaccine as part of the nationwide NHS vaccination campaign to rates for slightly younger individuals with the same characteristics who became eligible for vaccination later. Emergency admission was 50.1% (19.9% to 69.5%) less likely 21 to 27 days after vaccination and 75.6% (52.8% to 87.6%) less likely 35 to 41 days after first vaccination and 7 days after 80% had received their second dose. Covid-19 infection was 55.2% (40.8% to 66.8%) less likely 21 to 27 days after vaccination and 70.1% (55.1% to 80.1%) less likely 35 to 41 days after first vaccination and 7 days after 80% had received their second dose. Collectively these results are consistent with one dose of the BNT162b2 mRNA vaccine reducing events from 14 days after vaccination, with more effectiveness in reducing the severity of symptoms than preventing infection.

### Strengths and weaknesses of the study

We focused on a large number of the oldest people at high risk of serious Covid-19 outcomes. We considered a period and country experiencing widespread transmission and large numbers of hospitalisations. This provided statistical precision in the effectiveness estimates within a short period. We exploited a precise age cut-off that determined access to the vaccine, which reduced bias from selection into treatment.

Nonetheless, there is a risk of bias from unmeasured confounding. We matched cases and controls on combinations of 12 personal, household and area variables. We also compared four measures of hospital use in the previous 18 months and history of negative SARS-CoV-2 tests (see Appendix 6). Cases did not have lower event rates and had higher use of hospital services and more community-based Covid-19 tests prior to vaccination. This likely reflects the age difference which may bias our estimates towards lower than true effectiveness.

The rich set of matching variables meant some cases were excluded because there was no control available. These exclusions were more likely for some populations, including minority ethnic groups and residents of London, but the included individuals had similar outcomes to the excluded individuals and the effectiveness results were similar when we matched on fewer variables (Appendix 4).

The speed of the rollout of the NHS vaccination programme in England into younger populations reduced the pool of similar people who had not been vaccinated. We adjusted for the selection bias this generated and assessed the robustness of this adjustment by comparing to a younger age group where the selection bias occurred later in the monitoring period.

Finally, we considered Covid-19 related hospitalisations as well as positive Covid-19 tests. Hospitalisations are less likely to be influenced by changes in attitudes after receiving a vaccine that may affect whether individuals seek Covid-19 tests, such as misperceptions of immunity or misinterpretations of symptoms as side effects.

### Strengths and weaknesses in relation to other studies, discussing important differences in results

Our results are broadly consistent with existing estimates of BNT162b vaccine effectiveness, despite variations in study design, participant demographics and outcome definitions. We estimate effectiveness against documented infection of approximately 55% after one dose, rising to 70% after the majority received a second dose. These estimates are relatively consistent with results from a similar study in England (55% after one dose, 80% 7 days after all received a second dose),^8^ for the older age group in a similar study in Israel (50% after one dose, 95% 7 days after all received a second dose),^5^ and all-age results from an RCT (52% rising to 95%).^3^ Our point estimate of effectiveness against hospitalisation with Covid-19 (76% 7 days after most received a second dose) is somewhat lower than, though statistically compatible with, other estimates (80-87%)^5,8^.

Several factors likely contribute to these differences. First, our estimates are specifically for an older population where vaccine-induced immune responses may be sub-optimal^11^. In addition, our population included 20% of vaccinated individuals for whom the second dose was extended beyond the study period. This may explain the greater agreement with existing estimates for effectiveness 21-27 days after vaccination than for longer follow-up when second dose coverage varied between studies. Consistent with this, a study based in Scotland where the majority received only single dose BNT162b estimated 68% (53 to 79) effectiveness against hospitalisation at an equivalent timepoint (35-41 days post vaccination)^7^, slightly lower than our estimate.

Finally, an important consideration in observational studies is bias in selection into the intervention group. While all existing studies used statistical methods to adjust for biases,^5,7,8^ we exploited the precise age thresholds that determined temporal eligibility for vaccination, thereby reducing the risk of unmeasured confounding between cases and controls. Such biases are exacerbated with longer follow-up periods as those remaining unvaccinated become increasingly different from those vaccinated earlier. The divergence between our effectiveness estimates and those in other studies with longer follow-up may reflect less bias in our study design and adjustment methodology.

### Meaning of the study: possible explanations and implications for clinicians and policymakers

We provide evidence of high real-world effectiveness of the original dosing schedule of the BNT162b2 mRNA Covid-19 vaccine in preventing infections and hospitalisations despite the widespread transmission of the B.1.1.7 variant shortly after the study population was vaccinated. There have been concerns about reduced vaccine effectiveness, though our data is consistent with mass vaccination data^5,7,8^ and only slightly reduced neutralisation of B.1.1.7 pseudovirus relative to the Wuhan reference strain.^20^

### Unanswered questions and future research

Our study provides rigorous evidence to support effectiveness of vaccination in the real-world amongst older people. Future research priorities include the optimal dosing regimen, the longevity of this protection and applicability to other variants, effectiveness amongst younger people and specific population subgroups, and effects on onward transmission and asymptomatic infection.

## Data Availability

Data used in the study are not publicly available and may be obtained via NHS Digital (https://digital.nhs.uk/services/data-access-request-service-dars). The analysis scripts used for the evaluation are available at https://github.com/NHSEI-Analytics/nhs_covid19_effectiveness.

## Ethics

Surveillance of COVID-19 testing and vaccination is undertaken under Regulation 3 of The Health Service (Control of Patient Information) Regulations 2002 to collect confidential patient information (http://www.legislation.gov.uk/uksi/2002/1438/regulation/3/made) under Sections 3(i) (a) to (c), 3(i)(d) (i) and (ii) and 3(3). Review by the Heath Research Authority identified no regulatory issues with this evaluation, and ethical review is not a requirement for this study.

Data analysis was facilitated under Control of patient information (COPI) notice (https://digital.nhs.uk/coronavirus/coronavirus-covid-19-response-information-governance-hub/control-of-patient-information-copi-notice). Informed consent was not required for the study.

## Author contributions

TM conceived the idea and wrote the first draft of the manuscript. MW, JH and OA undertook the analysis. All authors contributed to the study design and the writing of the final manuscript. TM is the guarantor.

The corresponding author attests that all listed authors meet authorship criteria and that no others meeting the criteria have been omitted. TM affirms that the manuscript is an honest, accurate, and transparent account of the study being reported; that no important aspects of the study have been omitted; and that any discrepancies from the study as planned (and, if relevant, registered) have been explained.

## Copyright/license for publication

The Corresponding Author has the right to grant on behalf of all authors and does grant on behalf of all authors, a worldwide licence to the Publishers and its licensees in perpetuity, in all forms, formats and media (whether known now or created in the future), to i) publish, reproduce, distribute, display and store the Contribution, ii) translate the Contribution into other languages, create adaptations, reprints, include within collections and create summaries, extracts and/or, abstracts of the Contribution, iii) create any other derivative work(s) based on the Contribution, iv) to exploit all subsidiary rights in the Contribution, v) the inclusion of electronic links from the Contribution to third party material where-ever it may be located; and, vi) licence any third party to do any or all of the above.

## Public involvement

It was not appropriate or possible to involve patients or the public in the design, or conduct, or reporting, or dissemination plans of our research.

## Acknowledgements

We would like to acknowledge NHS England, Arden and GEM CSU, NHS Digital and NHS Test and Trace for their roles in developing and managing the various datasets used as part of the study. We are grateful to James Lockyer for developing the Master Patient Index, and to Stephen Smith, Nathan Abbots, Chris Jones and Natalie Talbott for their endeavours supplying the data in a form required for the analysis. We thank Kerrison Noonan for help collating the results of the study. We also thank Chris Gibbins, Rob Shaw, Seema Patel, Ed Kendall, Svetlana Batrakova and Simon Slovik for their advice and guidance, and Andrew Jackson for his continued support throughout.

## Supplementary Appendices

## Appendix 1 Data sources

NHS England & Improvement has access to person-level datasets for the entire population of England that provide information on who has been vaccinated, by date, vaccine type and dose, their age, gender and details of the address they live at, if they have had a prior COVID-19 infection (that is reported) and whether they go on to test positive for COVID-19 post vaccination. These datasets, that are collated by NHS Digital, Public Health England and the Office for National Statistics, include information on the contacts individuals had with the health service before and since being vaccinated, including A&E attendances and hospital admissions, and information regarding people who have since died. In combination, these datasets offer significant insights into what happens to people post their first vaccination dose to understand the effectiveness of the vaccines.

A Master Patient Index (MPI) data mart has been developed that includes details of all NHS registered patients in England. The MPI is built from extracts from the National Health Application and Infrastructure Services (NHAIS) system^1^, and comprises a list of NHS registered patients including details of their gender, age, area of residence together with a variety of derived data items including whether they are a permanent resident of a care home (based on address matching to registered care home details from the Care Quality Commission), detail of house occupancy/living arrangements (of which three categories have been used for the pairwise matching methodology: living with children under 18 years, living alone, and all other living arrangements) and supplementary information sourced from the Indices of Multiple Deprivation (IMD) classification for 2019.^2^

Several data assets have been linked to the MPI based on NHS Number (using a common pseudonym within a secure environment in accordance with the Control of Patient Information (COPI) notice^3^) including:

- Vaccinations event data sourced from the National Immunisation Management Service (NIMS)^4^ to provide details of the 1st and 2nd doses of the BNT162b2 mRNA and the ChAdOx1 adenovirus vector vaccines at person level. The extracts taken from the NIMS system undergo a series of transformations and data cleaning steps to identify events where vaccinations were given in line with those used for publication^5^. The dataset is also used to identify people who received a seasonal flu vaccine during FY2020/21;
- Hospital-based (Pillar 1) and community-based (Pillar 2) COVID-19 positive and negative tests data sourced from Public Health England Unified Sample Dataset^6^. For this analysis a dataset prepared by PHE that provides details of the 1st positive polymerase-chain-reaction (PCR) COVID-19 test per individual is used;
- Death registrations sourced from the Office for National Statistics (ONS)^7^;
- Accident and Emergency (A&E) attendance records sourced from the Emergency Care Dataset (ECDS) via NHS Digital^8^;
- Admitted patient care hospitals spell records sourced from the Admitted Patient Care Commissioning Dataset (APC CDS)^9^ via NHS Digital’s SUS+ Service^10^; and
- Shielded Patient List as generated by NHS Digital using a series of patient level collections to identify clinically extremely vulnerable individuals.^11^

A number of derived data items have been incorporated into the data mart to provide information on the health status of individuals, including the ‘Bridges to Health’ segmentation model developed by Outcomes Based Healthcare^12^ that identifies patients with co-morbidities that increase their risk of hospitalisation (here referred to as clinically vulnerable), and an algorithm applied to historic APC CDS spells data to identify frail individuals^13^. For the pairwise matching methodology, these data items have been combined into a measure of health status comprising three categories: frail and/or clinically extremely vulnerable, clinically vulnerable, and other/unknown (which predominately represents relatively healthy individuals).

Ethnicity data sourced from the ‘Bridged to Health’ model has been supplemented by data from NHS Digital sourced from a range of administrative sources include General Practice records. For the pairwise matching methodology four categories have been used: White, Black, Pakistani or Bangladeshi, other ethnicity, and ethnicity not reported. These categories are correlated with vaccine uptake in the wider population^14^.

A weighted acute illness measure has been derived using completed APC CDS spells. Total non-elective occupied bed days for individuals discharged between 15^th^ November and 14^th^ December have been multiplied by two, and the equivalent measure for discharges between the 15^th^ October 2020 and 14^th^ November 2020 remain unadjusted, with the two values summed. Spells where an individual was admitted and discharged on the same day count as 0.5 bed days. The acute illness measure is then defined as follows: high non-elective bed use (6 plus weighted bed days); medium non-elective bed use (3 to 5 weighted bed days), and low/no non-elective bed use (0 to 2 weighted bed days).

The refresh cycle for the data sources used in this data mart are daily for the vaccinations, testing, mortality, A&E and APC datasets. The MPI is refreshed on a monthly cycle. The coverage, completeness and quality of these collections vary, and steps have been taken to ensure the data used in the evaluation are complete. The data mart used to generate the results presented in this analysis was extracted on the 9^th^ February 2021 and includes complete records to the 3^rd^ February 2021. This uses a cut of the MPI from mid-November 2020 and all age-based calculations reference individuals ages as of mid-November 2020.

## Appendix 2 Matching and adjustment methodology

We matched vaccinated individuals in their early 80s to controls in their late 70s. This exploits the age-based eligibility criteria for the nationwide population vaccination^15^, whereby people aged 80 years and over were prioritised for vaccination. Whilst this approach helps minimise these biases in the early stages of the vaccination programme, from mid-January 2021 significant numbers of 70-year-olds had received their 1^st^ vaccine dose. Because we exclude individuals who were vaccinated more than 14 days before the end of the monitoring period from the pool of potential controls, this generates bias whereby the proportion of people in the pool available for matching becomes enriched in people who test positive for COVID-19 as individuals should not have had a COVID-19 infection in the two weeks prior to vaccination. If unaccounted for, this selection bias artificially increases the number of COVID-19 positive people in the pairwise control relative to the vaccinated cohort (see Table A2-1).

**Table A2-1.** Summary of change in the number of people and COVID-19 status of people in the available for matching to vaccinated individuals as the monitoring period used in the evaluation is extended. The analysis has been run twice: once excluding controls that received their 1^st^ COVID-19 dose during the monitoring period, and a second time excluding controls that received their 1^st^ COVID-19 dose before 14 days of the end of the monitoring period (which is the approach used in the main analysis).

| Days since vaccination | Total number of 80 to 83 year olds vaccinated | Pairwise matching excluding controls that received their 1st COVID-19 vaccine dose during the monitoring period |  |  |  |  | Pairwise matching excluding controls that received their 1st COVID-19 vaccine dose before 14 days of the end of the monitoring period |  |  |  |  |
| --- | --- | --- | --- | --- | --- | --- | --- | --- | --- | --- | --- |
|  |  | Total number of match pairs | % Match rate | Count of potential controls | Count of potential controls with a positive COVID-19 test result | Percentage of potential controls with a positive COVID-19 test result | Total number of match pairs | % Match rate | Count of potential controls | Count of potential controls with a positive COVID-19 test result | Percentage of potential controls with a positive COVID-19 test result |
| Day 11 | 170,424 | 129,939 | 76.2% | 1,929,275 | 23,857 | 1.2% | 131,236 | 77.0% | 1,949,529 | 24,272 | 1.2% |
| Day 12 | 170,424 | 129,742 | 76.1% | 1,926,965 | 23,823 | 1.2% | 131,143 | 77.0% | 1,948,512 | 24,262 | 1.2% |
| Day 13 | 170,424 | 129,519 | 76.0% | 1,924,717 | 23,801 | 1.2% | 131,031 | 76.9% | 1,947,080 | 24,254 | 1.2% |
| Day 14 | 170,424 | 129,523 | 76.0% | 1,924,321 | 23,796 | 1.2% | 130,960 | 76.8% | 1,945,651 | 24,242 | 1.2% |
| Day 15 | 170,424 | 129,474 | 76.0% | 1,924,136 | 23,793 | 1.2% | 130,816 | 76.8% | 1,944,085 | 24,218 | 1.2% |
| Day 16 | 170,424 | 129,364 | 75.9% | 1,922,923 | 23,777 | 1.2% | 130,782 | 76.7% | 1,942,649 | 24,207 | 1.2% |
| Day 17 | 170,424 | 129,116 | 75.8% | 1,918,750 | 23,723 | 1.2% | 130,736 | 76.7% | 1,941,076 | 24,192 | 1.2% |
| Day 18 | 170,424 | 128,298 | 75.3% | 1,899,266 | 23,531 | 1.2% | 130,709 | 76.7% | 1,940,620 | 24,173 | 1.2% |
| Day 19 | 170,424 | 127,295 | 74.7% | 1,873,909 | 23,297 | 1.2% | 130,695 | 76.7% | 1,940,618 | 24,173 | 1.2% |
| Day 20 | 170,424 | 126,223 | 74.1% | 1,854,501 | 23,118 | 1.2% | 130,703 | 76.7% | 1,940,612 | 24,173 | 1.2% |
| Day 21 | 170,424 | 125,200 | 73.5% | 1,845,495 | 23,029 | 1.2% | 130,671 | 76.7% | 1,940,600 | 24,173 | 1.2% |
| Day 22 | 170,424 | 124,613 | 73.1% | 1,838,815 | 22,924 | 1.2% | 130,709 | 76.7% | 1,940,548 | 24,173 | 1.2% |
| Day 23 | 170,424 | 123,842 | 72.7% | 1,826,997 | 22,815 | 1.2% | 130,667 | 76.7% | 1,939,679 | 24,138 | 1.2% |
| Day 24 | 170,424 | 122,749 | 72.0% | 1,796,769 | 22,549 | 1.3% | 130,293 | 76.5% | 1,934,746 | 23,971 | 1.2% |
| Day 25 | 170,424 | 120,994 | 71.0% | 1,762,271 | 22,281 | 1.3% | 129,880 | 76.2% | 1,929,275 | 23,857 | 1.2% |
| Day 26 | 170,424 | 118,830 | 69.7% | 1,720,682 | 21,961 | 1.3% | 129,687 | 76.1% | 1,926,965 | 23,823 | 1.2% |
| Day 27 | 170,424 | 116,564 | 68.4% | 1,685,512 | 21,717 | 1.3% | 129,516 | 76.0% | 1,924,717 | 23,801 | 1.2% |
| Day 28 | 170,424 | 115,625 | 67.8% | 1,670,879 | 21,604 | 1.3% | 129,473 | 76.0% | 1,924,321 | 23,796 | 1.2% |
| Day 29 | 170,424 | 114,915 | 67.4% | 1,658,826 | 21,495 | 1.3% | 129,487 | 76.0% | 1,924,136 | 23,793 | 1.2% |
| Day 30 | 170,424 | 112,743 | 66.2% | 1,603,655 | 21,166 | 1.3% | 129,371 | 75.9% | 1,922,923 | 23,777 | 1.2% |
| Day 31 | 170,424 | 109,937 | 64.5% | 1,540,031 | 20,856 | 1.4% | 129,098 | 75.8% | 1,918,750 | 23,723 | 1.2% |
| Day 32 | 170,424 | 104,142 | 61.1% | 1,456,681 | 20,405 | 1.4% | 128,225 | 75.2% | 1,899,266 | 23,531 | 1.2% |
| Day 33 | 170,424 | 96,619 | 56.7% | 1,340,317 | 19,787 | 1.5% | 127,282 | 74.7% | 1,873,909 | 23,297 | 1.2% |
| Day 34 | 170,424 | 86,486 | 50.7% | 1,213,893 | 19,224 | 1.6% | 126,204 | 74.1% | 1,854,501 | 23,118 | 1.2% |
| Day 35 | 170,424 | 83,476 | 49.0% | 1,171,084 | 18,994 | 1.6% | 125,156 | 73.4% | 1,845,495 | 23,029 | 1.2% |
| Day 36 | 170,424 | 80,657 | 47.3% | 1,132,764 | 18,774 | 1.7% | 124,590 | 73.1% | 1,838,815 | 22,924 | 1.2% |
| Day 37 | 170,424 | 77,236 | 45.3% | 1,089,302 | 18,521 | 1.7% | 123,798 | 72.6% | 1,826,997 | 22,815 | 1.2% |
| Day 38 | 170,424 | 74,571 | 43.8% | 1,049,609 | 18,255 | 1.7% | 122,684 | 72.0% | 1,796,769 | 22,549 | 1.3% |
| Day 39 | 170,424 | 69,833 | 41.0% | 976,628 | 17,826 | 1.8% | 120,955 | 71.0% | 1,762,271 | 22,281 | 1.3% |
| Day 40 | 170,424 | 58,354 | 34.2% | 858,984 | 17,189 | 2.0% | 118,804 | 69.7% | 1,720,682 | 21,961 | 1.3% |
| Day 41 | 170,424 | 46,962 | 27.6% | 762,004 | 16,610 | 2.2% | 116,539 | 68.4% | 1,685,512 | 21,717 | 1.3% |
| Day 42 | 170,424 | 41,695 | 24.5% | 714,711 | 16,285 | 2.3% | 115,584 | 67.8% | 1,670,879 | 21,604 | 1.3% |
| Day 43 | 170,424 | 37,394 | 21.9% | 675,280 | 15,990 | 2.4% | 114,908 | 67.4% | 1,658,826 | 21,495 | 1.3% |
| Day 44 | 170,424 | 33,956 | 19.9% | 645,526 | 15,663 | 2.4% | 112,733 | 66.1% | 1,603,655 | 21,166 | 1.3% |
| Day 45 | 170,424 | 31,200 | 18.3% | 620,896 | 15,366 | 2.5% | 109,906 | 64.5% | 1,540,031 | 20,856 | 1.4% |

To adjust for this bias, we have developed a methodology where we generate a number of daily timeseries for each outcome by extending the monitoring period a day at a time, starting with an end date 11 days post vaccination (at which point the bias is minimal) and repeating the process 35 times to extend the monitoring period to 45 days post vaccination. With each iteration the size of the potential pool of match pairs contracts as we exclude from the pool of potential controls individuals who were vaccinated more than 14 days before the end of the monitoring period.

We then align each timeseries so that vaccination events on the 15^th^, 16^th^, 17^th^, 18^th^, 19^th^ and 20^th^ December 2020 reference to day 0 with up to 45 days follow-up. The absolute numbers for each outcome (positive tests, A&E attendances with COVID-19 and admissions with COVID-19 via A&E) to hospital) are converted into rate per 100,000 per day by dividing by the size of the matched cohort for each of the 35 runs.

Next we compare the total cumulative rate for each run to the previous day’s run excluding the latest day from the former to calculate the change run-on-run (a_d_) as below:

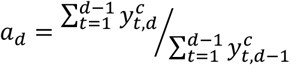

where 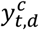s the event rate per 100,000 persons at time t for the control group, c represents the control cohort, *t* is the start date of the monitoring period (which is fix as Day 1), and *d* is the last day for each run. This generates a set of values for *a*_*d*_ that are applied to correct for the cumulative sampling bias using as follows:

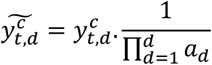

Following these steps, we generate a set of adjusted time series for each run for the controls and vaccinated cohorts as illustrated in Figure A2-1. For each run we take the latest value as the best estimate of the adjusted daily figure for that point in time as denoted by the dashed line.

**Figure A2-1.**
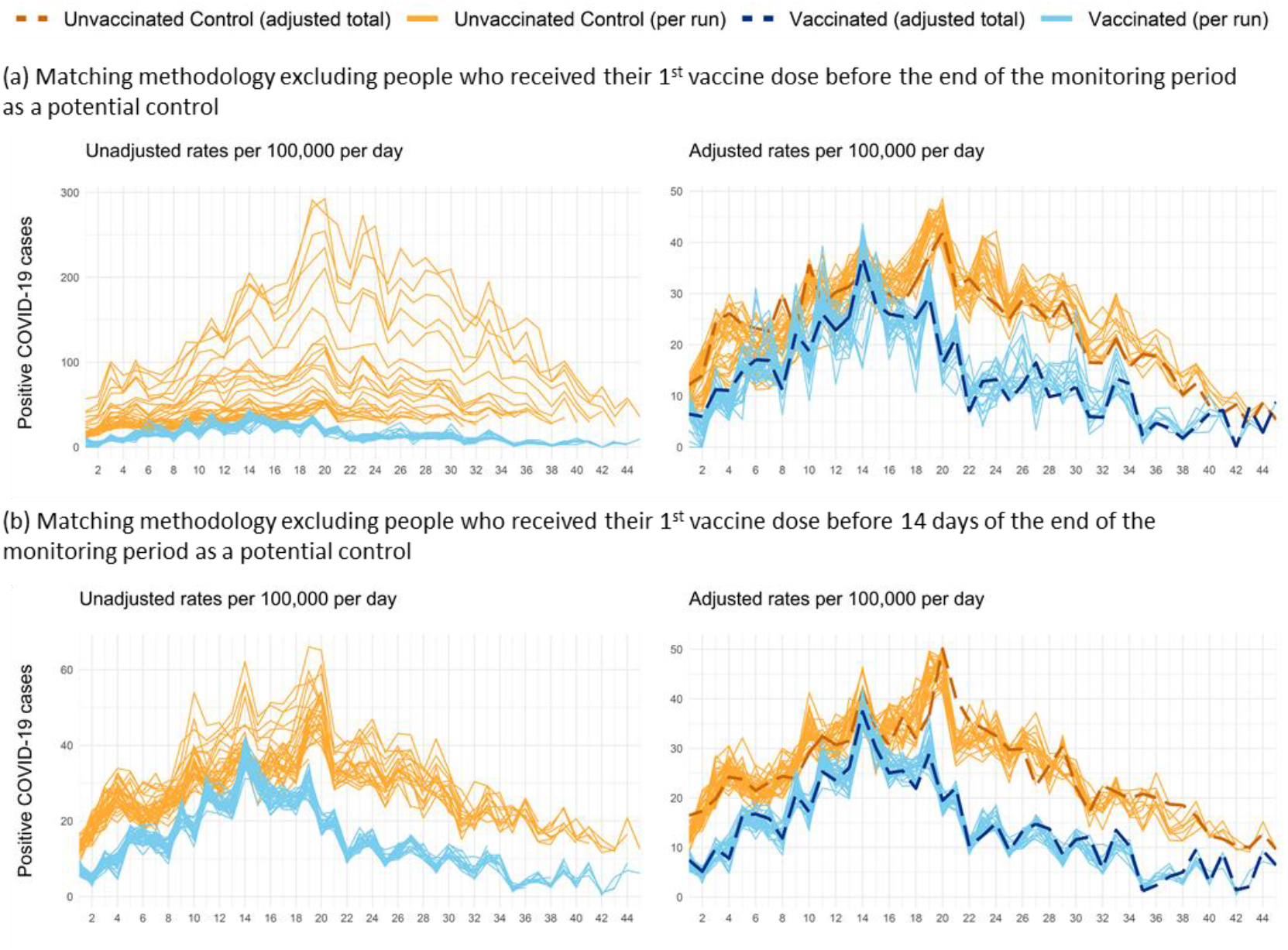
Numbers of individuals testing positive for COVID-19 post vaccination (in blue) with a comparison to their match pairs (in orange) as a rate per 100,000. The left-hand charts present the raw numbers pre-adjustment for the sampling bias where each line represents to a different cut-off date for the analysis that varies the size of the pool of individuals available for matching. The right-hand charts present the adjusted figures based on the methodology, where the dashed blue and dashed orange lines represent the most complete estimate by day since vaccination.

The process is repeated five times for five separate batches (where the matching process is repeated with different random number seeds, allowing replacement in match-pairs between the sensitivity runs). A simple average is taken across the five batches as follows:

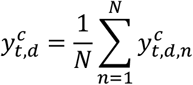

We then run a bootstrapping procedure with replacement 100 times within each of the batches. This gives us 5×100 values for each of the key statistics (event rate in vaccinated, event rate in matched controls, relative reduction in event rate). We use the distribution of these statistics to generate 95% confidence intervals (by picking the 12th (0.025*500) lowest and highest values). For graphical purposes only, seven-day moving averages of the rates are calculated and presented.

## Appendix 3 Measuring COVID-19 related emergency hospital attendances and admissions

Two outcome measures of hospitalisation with COVID-19 have been used in the evaluation: (1) A&E attendance with Covid-19 measured using the Emergency Care Data Set (ECDS), and (2) non-elective admission to hospital measured using the Admitted Patient Care Commissioning Dataset (APC CDS). Both datasets have timeliness issues with coverage and coding completeness that can bias analysis if not accounted for.

To circumvent issues with coding completeness, a matching algorithm has been used to identify COVID-19 related A&E attendances and admissions by combining diagnosis information available from the ECDS/APC records (see Table A3-1) with COVID-19 positive test results where the specimen was taken between 14 before and 6 days after the linked A&E attendance/admission. Using this matching window, 23% of linked test results were taken between 14 and 1 days before the A&E attendance, with 58% having a linked tests taken on days 0 to 1 post attendance, and 19% having a linked test taken between days 2 and 6 days post attendance (see Figure A3-1). Coding completeness is less of an issue with the APC CDS data, where the majority of COVID-19 related admissions both have a COVID-19 diagnosis on the APC record, and a linked COVID-19 positive test result (see Figure A3-2).

**Table A3-1.** List of SNOMED CT and ICD-10 codes used to identify 1^st^ COVID-19 related A&E attendances and hospital admissions based on the primary diagnosis, secondary diagnoses, and (for ECDS) notifiable diseases fields within the ECDS and APC CDS collections. Several additional SNOMED CT codes are available for certain conditions that present with COVID-19, but an analysis suggest none of these codes (and other associated codes for COVID-19) have been used within the ECDS collection.

| SNOMED CT Code/<br>ICD-10 Code | Description |
| --- | --- |
| 1240751000000100 | Coronavirus disease 19 caused by severe acute respiratory syndrome coronavirus 2 (disorder) |
| 1240761000000102 | Suspected coronavirus disease 19 caused by severe acute respiratory syndrome coronavirus 2 (situation) |
| 1300721000000109 | Coronavirus disease 19 caused by severe acute respiratory syndrome coronavirus 2 confirmed by laboratory test (situation) |
| U07.1 | COVID-19, virus identified |
| U07.2 | COVID-19, virus not identified |
| B97.2 | Coronavirus as the cause of diseases classified to other chapters |

**Figure A3-1.**
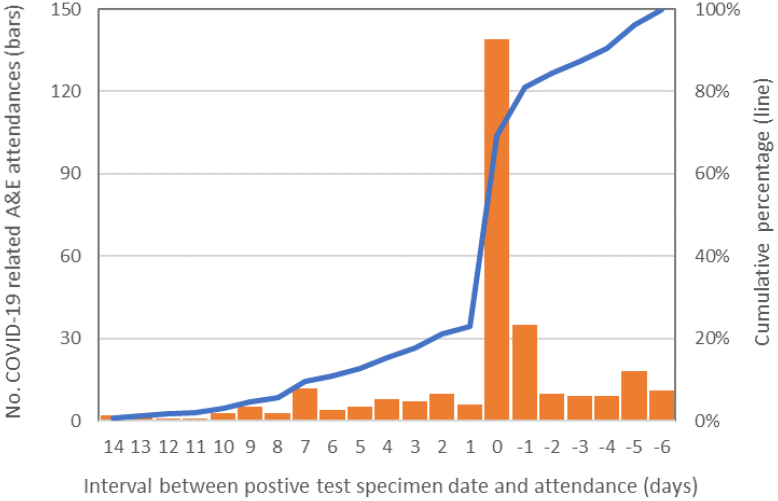
Delay between specimen data for a COVID-19 positive test and the associated A&E attendance for 80-to 83-year-olds vaccinated between the 15^th^ and 20^th^ December 2020.

**Figure A3-2.**
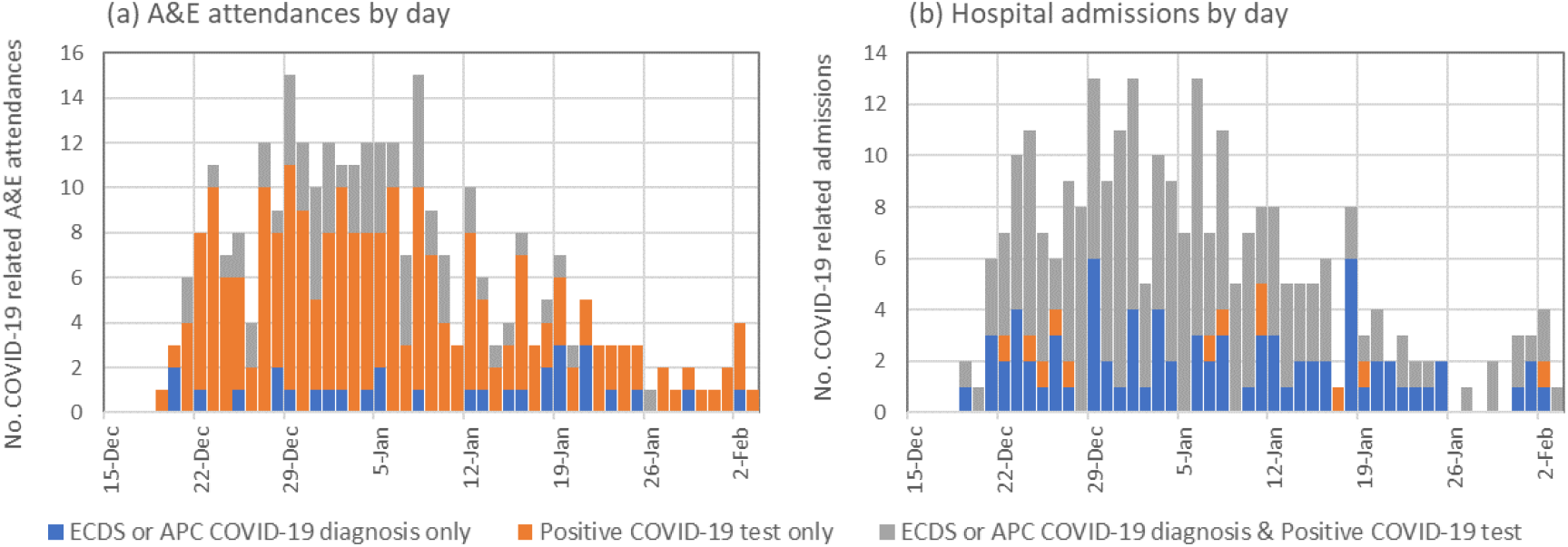
Counts of COVID-19 related A&E attendances and admissions for 80-to 83-year-olds vaccinated between the 15^th^ and 20^th^ December 2020 by method of identification, for (a) attendances, and (b) admissions.

The matching algorithm allows all A&E attendances and/or admissions within this matching window to be counted as COVID-19 related, or it can be constrained to select the first COVID-19 related attendance and/or admission per person. Using the latter approach avoids counting multiple attendances and/or admissions for the same person and is the approach used in this analysis.

For the ACP CDS, an additional complication relates to the collection being discharge centric whereby providers usually only submit records for patients that has been discharged from hospital. To allow reporting by admission date, we restricted the analysis to patients who stayed in hospital 42 days or less. This excludes 5.7% of COVID-19 related spells for 80-to 83-year olds (based on patients discharged between the 8^th^ December 2020 and the 31^st^ March 2021).

To account for incomplete coverage of the ECDS collection, the analysis is based on A&E data for a sample of 118 NHS providers in England that have complete data to the 9^th^ February 2021 based on an extract taken on the 9^th^ March 2021. Similarly, to account for incomplete coverage of the APC CDS collection, the analysis is based on APC CDS data for a sample of 124 NHS providers in England that have complete data to the 17^th^ March 2021 based on an extract taken on the 6^th^ April 2021. These two samples accounts for 95% of Type 1&2 A&E attendances and 93% of admitted patient care spell for England. As the pairwise control matching methodology matches the vaccinated individual to a control from the same middle super output area, we do not expect a systematic bias in the analysis due to incomplete coverage.

These attendances and admissions are referred to as ‘with’ COVID-19 as the available data does not allow us to definitively determine if their COVID-19 infection was the primary reason for their A&E attendance or admission to hospital.

## Appendix 4 Changing composition of the study population by follow-up period

The adjustment methodology described in Appendix 2 involves re-matching vaccinated individuals to controls as the follow-up period is extended a day at a time. Therefore, depending on the length of follow-up, the size and composition of both the vaccinated group and unvaccinated control group change. For example, in the main analysis, the match rate falls from 77.0% at day 11 (which represents the baseline case) to 64.5% at day 45 (Table A2-1).

Table A4-1 presents the number and percentage of vaccinated individuals for whom a pairwise match was identified at the mid-points of each period of follow-up, stratified by each matching variable. While there is wide variability in the percentage of vaccinated individuals with a matched pair (from 1.2% to 84.2%) across subgroups at day 17 of follow-up, the relative composition of those included in analysis is relatively stable across extended follow-up times (Table A4-1). The largest changes were for health status category where the share of the health/other category increases by 0.41 percentage points, with concurrent reductions of 0.22 and 0.18 percentage points for the frail and/or clinically extremely vulnerable and clinical vulnerable groups respectively between 17 and 38. These changes are small and are unlikely to result in any significant bias to the effectiveness estimates as the monitoring period is extended.

Table A4-1 also provides unadjusted and adjusted estimates of cumulative documented infections amongst vaccinated and unvaccinated control individuals by subgroup. These were generated by selecting all records without replacement and running the same adjustment process that was used in the bootstrapping process in the main analysis. Note that the main analysis bootstrapping process sampled with replacement, so values differ from those in the main analysis. We derived the ratio of adjusted to unadjusted total infections and applied this as a normalisation factor to each subgroup to estimate adjusted subgroup counts.

**Table A4-1.** Demographic and clinical characteristics of vaccinated persons and their unvaccinated controls based on matching to the 76 to 79 years population. Four matched cohort at days 17, 24, 31 and 38 post vaccination are presented, which relate to the mid-points of each period of follow-up. Cumulative unadjusted and adjusted rates of COVID-19 positive tests are included to demonstrate differences in test-positivity rates between the subgroups.

|  |  |  |  | Days after vaccination<br>17 |  |  |  |  |  | Days after vaccination<br>24 |  |  |  |  |  |  |  |  |
| --- | --- | --- | --- | --- | --- | --- | --- | --- | --- | --- | --- | --- | --- | --- | --- | --- | --- | --- |
| Matching criteria | Categories | Total Number of<br>80 to 83 year<br>olds | Of which tested<br>positive for<br>COVID-19 by<br>day 45 per<br>100,000 | Number<br>with a<br>pairwise<br>control<br>match | % with a<br>pairwise<br>match | % of total<br>match pairs | Vaccinated -<br>tested positive<br>for COVID per<br>100,000<br>(unadjusted) | Vaccinated -<br>tested positive<br>for COVID per<br>100,000<br>(adjusted) | Unvaccinated -<br>tested positive<br>for COVID per<br>100,000<br>(unadjusted) | Unvaccinated -<br>tested positive<br>for COVID per<br>100,000<br>(adjusted) | Number<br>with a<br>pairwise<br>control<br>match | % with a<br>pairwise<br>match | % of total<br>match pairs | Vaccinated -<br>tested<br>positive for<br>COVID<br>(unadjusted) | Vaccinated -<br>tested<br>positive for<br>COVID per<br>100,000<br>(unadjusted) | Vaccinated -<br>tested<br>positive for<br>COVID per<br>100,000<br>(adjusted) | Unvaccinated -<br>tested<br>positive for<br>COVID per<br>100,000<br>(unadjusted) | Unvaccinated -<br>tested<br>positive for<br>COVID per<br>100,000<br>(adjusted) |
| Gender | Female | 89,930 | 713 | 69,566 | 77.4% | 53.2% | 346.4 | 344.4 | 474.4 | 443.3 | 69,323 | 77.1% | 53.2% | 316 | 455.8 | 460.2 | 680.9 | 665.1 |
|  | Male | 80,295 | 757 | 61,170 | 76.2% | 46.8% | 317.1 | 315.3 | 439.8 | 411.0 | 60,970 | 75.9% | 46.8% | 284 | 465.8 | 470.2 | 683.9 | 668.1 |
|  | Unknown | <5 | - | - | 0.0% | 0.0% | - | - | - | - | - | 0.0% | 0.0% | - | - | - | - | - |
| Ethnicity | White | 157,896 | 690 | 127,137 | 80.5% | 97.2% | 325.6 | 323.7 | 449.9 | 420.5 | 126,706 | 80.2% | 97.2% | 568 | 448.3 | 452.6 | 668.5 | 653.0 |
|  | Black | 1,567 | 1,595 | 270 | 17.2% | 0.2% | - | - | - | - | 270 | 17.2% | 0.2% | 3 | 1,111.1 | - | 1,111.1 | - |
|  | Pakistani and Bangladeshi | 1,141 | 3,243 | 210 | 18.4% | 0.2% | 2,381.0 | 2,367.1 | - | - | 210 | 18.4% | 0.2% | 6 | 2,857.1 | 2,884.4 | 2,857.1 | 2,790.9 |
|  | Other BAME | 5,922 | 1,418 | 1,778 | 30.0% | 1.4% | 731.2 | 726.9 | 1,181.1 | 1,103.8 | 1,769 | 29.9% | 1.4% | 20 | 1,130.6 | 1,141.4 | 1,469.8 | 1,435.7 |
|  | Unknown | 3,700 | 378 | 1,341 | 36.2% | 1.0% | - | - | - | - | 1,338 | 36.2% | 1.0% | 3 | 224.2 | - | 523.2 | 511.0 |
| IMD Quintile | 1 (most deprived) | 23,292 | 992 | 17,045 | 73.2% | 13.0% | 416.5 | 414.1 | 580.8 | 542.8 | 16,993 | 73.0% | 13.0% | 98 | 576.7 | 582.2 | 847.4 | 827.8 |
|  | 2 | 28,556 | 823 | 20,013 | 70.1% | 15.3% | 374.8 | 372.6 | 584.6 | 546.4 | 19,956 | 69.9% | 15.3% | 104 | 521.1 | 526.1 | 761.7 | 744.0 |
|  | 3 | 35,785 | 819 | 26,801 | 74.9% | 20.5% | 328.3 | 326.4 | 421.6 | 394.0 | 26,744 | 74.7% | 20.5% | 128 | 478.6 | 483.2 | 676.8 | 661.1 |
|  | 4 | 40,339 | 642 | 31,564 | 78.2% | 24.1% | 297.8 | 296.1 | 434.0 | 405.6 | 31,427 | 77.9% | 24.1% | 127 | 404.1 | 408.0 | 661.9 | 646.5 |
|  | 5 (least deprived) | 42,213 | 545 | 35,313 | 83.7% | 27.0% | 303.0 | 301.2 | 376.6 | 352.0 | 35,173 | 83.3% | 27.0% | 143 | 406.6 | 410.4 | 580.0 | 566.5 |
|  | Not recorded | 41 | 2,439 | - | 0.0% | 0.0% | - | - | - | - | - | 0.0% | 0.0% | - | - | - | - | - |
| Living arrangements | Living with children | 5,297 | 1,793 | 1,209 | 22.8% | 0.9% | 661.7 | 657.8 | 744.4 | 695.7 | 1,204 | 22.7% | 0.9% | 12 | 996.7 | 1,006.2 | 1,578.1 | 1,541.5 |
|  | Living alone | 49,876 | 664 | 33,613 | 67.4% | 25.7% | 318.3 | 316.5 | 482.0 | 450.4 | 33,471 | 67.1% | 25.7% | 153 | 457.1 | 461.5 | 648.3 | 633.3 |
|  | All other living arrangements | 115,053 | 715 | 95,914 | 83.4% | 73.4% | 333.6 | 331.7 | 446.2 | 417.0 | 95,618 | 83.1% | 73.4% | 435 | 454.9 | 459.3 | 682.9 | 667.1 |
| Health Risk | Frail and/or clinically extremely vulnerable | 42,856 | 1,199 | 28,780 | 67.2% | 22.0% | 517.7 | 514.7 | 667.1 | 623.5 | 28,614 | 66.8% | 22.0% | 216 | 754.9 | 762.1 | 1,044.9 | 1,020.7 |
|  | Clinically vulnerable | 57,742 | 727 | 43,300 | 75.0% | 33.1% | 330.3 | 328.3 | 480.4 | 448.9 | 43,147 | 74.7% | 33.1% | 201 | 465.8 | 470.3 | 748.6 | 731.2 |
|  | Healthy/other | 69,628 | 452 | 58,656 | 84.2% | 44.9% | 243.8 | 242.4 | 339.3 | 317.1 | 58,532 | 84.1% | 44.9% | 183 | 312.6 | 315.6 | 456.2 | 445.6 |
| Flu vaccine | Received seasonal flu vaccine FY2020/21 | 150,300 | 710 | 118,292 | 78.7% | 90.5% | 323.8 | 321.9 | 467.5 | 436.9 | 117,906 | 78.4% | 90.5% | 530 | 449.5 | 453.8 | 681.9 | 666.1 |
|  | Did not received seasonal flu vaccine FY2020/22 | 19,926 | 913 | 12,444 | 62.5% | 9.5% | 417.9 | 415.4 | 369.7 | 345.5 | 12,387 | 62.2% | 9.5% | 70 | 565.1 | 570.5 | 686.2 | 670.3 |
| Recent acute illness | High | 809 | 3,214 | 22 | 2.7% | 0.0% | - | - | - | - | 22 | 2.7% | 0.0% | - | - | - | 4,545.5 | - |
|  | Medium | 521 | 3,647 | 6 | 1.2% | 0.0% | - | - | - | - | 6 | 1.2% | 0.0% | - | - | - | - | - |
|  | Low | 168,896 | 713 | 130,708 | 77.4% | 100.0% | 332.8 | 330.9 | 458.3 | 428.3 | 130,265 | 77.1% | 100.0% | 600 | 460.6 | 465.0 | 681.7 | 665.9 |
| Region | East Midlands | 8,830 | 657 | 7,131 | 80.8% | 5.5% | 252.4 | 250.9 | 350.6 | 327.6 | 7,115 | 80.6% | 5.5% | 29 | 407.6 | 411.5 | 646.5 | 631.5 |
|  | East of England | 21,805 | 807 | 17,276 | 79.2% | 13.2% | 445.7 | 443.1 | 497.8 | 465.2 | 17,262 | 79.2% | 13.2% | 101 | 585.1 | 590.7 | 724.1 | 707.3 |
|  | London | 18,923 | 1,575 | 10,804 | 57.1% | 8.3% | 814.5 | 809.8 | 1,055.2 | 986.1 | 10,708 | 56.6% | 8.2% | 102 | 952.6 | 961.7 | 1,512.9 | 1,477.8 |
|  | North East | 9,300 | 419 | 7,323 | 78.7% | 5.6% | 163.9 | 162.9 | 587.2 | 548.8 | 7,257 | 78.0% | 5.6% | 23 | 316.9 | 320.0 | 744.1 | 726.9 |
|  | North West | 22,898 | 751 | 18,200 | 79.5% | 13.9% | 340.7 | 338.7 | 450.5 | 421.1 | 18,145 | 79.2% | 13.9% | 93 | 512.5 | 517.4 | 760.5 | 742.9 |
|  | South East | 29,011 | 707 | 22,580 | 77.8% | 17.3% | 367.6 | 365.4 | 451.7 | 422.2 | 22,516 | 77.6% | 17.3% | 115 | 510.7 | 515.6 | 759.5 | 741.9 |
|  | South West | 21,349 | 436 | 17,290 | 81.0% | 13.2% | 156.2 | 155.2 | 214.0 | 200.0 | 17,190 | 80.5% | 13.2% | 45 | 261.8 | 264.3 | 337.4 | 329.6 |
|  | West Midlands | 22,168 | 623 | 17,623 | 79.5% | 13.5% | 221.3 | 220.0 | 408.6 | 381.8 | 17,596 | 79.4% | 13.5% | 57 | 323.9 | 327.0 | 500.1 | 488.5 |
|  | Yorkshire and The Humber | 15,882 | 434 | 12,509 | 78.8% | 9.6% | 231.8 | 230.5 | 303.8 | 283.9 | 12,504 | 78.7% | 9.6% | 35 | 279.9 | 282.6 | 375.9 | 367.2 |
|  | Unknown | 60 | - | - | 0.0% | 0.0% | - | - | - | - | - | 0.0% | 0.0% | - | - | - | - | - |
| Total |  | 170,226 | 733 | 130,736 | 76.8% | 100.0% | 332.7 | 330.8 | 458.2 | 428.2 | 130,293 | 76.5% | 100.0% | 600 | 460.5 | 464.9 | 682.3 | 666.5 |

**Table A4-1 (Cont.)** Demographic and clinical characteristics of vaccinated persons and their unvaccinated controls based on matching to the 76 to 79 years population.
|  |  |  |  | Days after vaccination |  |  |  |  |  | Days after vaccination |  |  |  |  |  |  |  |  |
| --- | --- | --- | --- | --- | --- | --- | --- | --- | --- | --- | --- | --- | --- | --- | --- | --- | --- | --- |
|  |  |  |  | 31 |  |  |  |  |  | 38 |  |  |  |  |  |  |  |  |
| Matching criteria | Categories | Total Number of<br>80 to 83 year<br>olds | Of which tested<br>positive for<br>COVID-19 by<br>day 45 per<br>100,000 | Number<br>with a<br>pairwise<br>control<br>match | % with a<br>pairwise<br>match | % of total<br>match pairs | Vaccinated -<br>tested positive<br>for COVID per<br>100,000<br>(unadjusted) | Vaccinated -<br>tested positive<br>for COVID per<br>100,000<br>(adjusted) | Unvaccinated -<br>tested positive<br>for COVID per<br>100,000<br>(unadjusted) | Unvaccinated -<br>tested positive<br>for COVID per<br>100,000<br>(adjusted) | Number<br>with a<br>pairwise<br>control<br>match | % with a<br>pairwise<br>match | % of total<br>match pairs | Vaccinated -<br>tested<br>positive for<br>COVID<br>(unadjusted) | Vaccinated -<br>tested<br>positive for<br>COVID per<br>100,000<br>(unadjusted) | Vaccinated -<br>tested<br>positive for<br>COVID per<br>100,000<br>(adjusted) | Unvaccinated -<br>tested<br>positive for<br>COVID per<br>100,000<br>(unadjusted) | Unvaccinated -<br>tested<br>positive for<br>COVID per<br>100,000<br>(adjusted) |
| Gender | Female | 89,930 | 713 | 68,684 | 76.4% | 53.2% | 531.4 | 530.3 | 861.9 | 824.6 | 65,285 | 72.6% | 53.2% | 370 | 566.7 | 565.6 | 1,066.1 | 953.4 |
|  | Male | 80,295 | 757 | 60,414 | 75.2% | 46.8% | 559.5 | 558.3 | 902.1 | 863.1 | 57,399 | 71.5% | 46.8% | 347 | 604.5 | 603.3 | 1,097.6 | 981.6 |
|  | Unknown | <5 | - | - | 0.0% | 0.0% |  |  |  |  | - | 0.0% | 0.0% | - |  |  |  |  |
| Ethnicity | White | 157,896 | 690 | 125,543 | 79.5% | 97.2% | 531.3 | 530.2 | 869.0 | 831.4 | 119,357 | 75.6% | 97.3% | 681 | 570.6 | 569.4 | 1,065.7 | 953.1 |
|  | Black | 1,567 | 1,595 | 270 | 17.2% | 0.2% | 2,222.2 | 2,217.7 | 1,111.1 |  | 253 | 16.1% | 0.2% | 6 | 2,371.5 | 2,366.6 | 1,185.8 |  |
|  | Pakistani and Bangladeshi | 1,141 | 3,243 | 210 | 18.4% | 0.2% | 2,857.1 | 2,851.3 | 2,857.1 | 2,733.6 | 188 | 16.5% | 0.2% | 7 | 3,723.4 | 3,715.7 | 3,191.5 | 2,854.2 |
|  | Other BAME | 5,922 | 1,418 | 1,749 | 29.5% | 1.4% | 1,200.7 | 1,198.2 | 1,715.3 | 1,641.1 | 1,632 | 27.6% | 1.3% | 20 | 1,225.5 | 1,222.9 | 2,328.4 | 2,082.3 |
|  | Unknown | 3,700 | 378 | 1,326 | 35.8% | 1.0% | 226.2 |  | 527.9 | 505.1 | 1,254 | 33.9% | 1.0% | 3 | 239.2 |  | 558.2 | 499.2 |
| IMD Quintile | 1 (most deprived) | 23,292 | 992 | 16,858 | 72.4% | 13.1% | 711.8 | 710.4 | 996.6 | 953.5 | 16,001 | 68.7% | 13.0% | 134 | 837.4 | 835.7 | 1,456.2 | 1,302.2 |
|  | 2 | 28,556 | 823 | 19,817 | 69.4% | 15.4% | 595.4 | 594.2 | 1,059.7 | 1,013.9 | 18,720 | 65.6% | 15.3% | 114 | 609.0 | 607.7 | 1,314.1 | 1,175.2 |
|  | 3 | 35,785 | 819 | 26,460 | 73.9% | 20.5% | 559.3 | 558.2 | 959.9 | 918.4 | 25,182 | 70.4% | 20.5% | 158 | 627.4 | 626.1 | 1,064.3 | 951.8 |
|  | 4 | 40,339 | 642 | 31,129 | 77.2% | 24.1% | 472.2 | 471.3 | 812.7 | 777.6 | 29,608 | 73.4% | 24.1% | 149 | 503.2 | 502.2 | 945.7 | 845.7 |
|  | 5 (least deprived) | 42,213 | 545 | 34,834 | 82.5% | 27.0% | 488.0 | 487.0 | 723.4 | 692.1 | 33,173 | 78.6% | 27.0% | 162 | 488.3 | 487.3 | 901.3 | 806.1 |
|  | Not recorded | 41 | 2,439 | - | 0.0% | 0.0% |  |  |  |  | - | 0.0% | 0.0% | - |  |  |  |  |
| Living arrangements | Living with children | 5,297 | 1,793 | 1,188 | 22.4% | 0.9% | 1,262.6 | 1,260.0 | 1,431.0 | 1,369.1 | 1,100 | 20.8% | 0.9% | 13 | 1,181.8 | 1,179.4 | 1,636.4 | 1,463.4 |
|  | Living alone | 49,876 | 664 | 33,115 | 66.4% | 25.7% | 504.3 | 503.3 | 839.5 | 803.2 | 31,250 | 62.7% | 25.5% | 177 | 566.4 | 565.2 | 982.4 | 878.6 |
|  | All other living arrangements | 115,053 | 715 | 94,795 | 82.4% | 73.4% | 549.6 | 548.5 | 888.2 | 849.8 | 90,334 | 78.5% | 73.6% | 527 | 583.4 | 582.2 | 1,108.1 | 991.0 |
| Health Risk | Frail and/or clinically extremely vulnerable | 42,856 | 1,199 | 28,283 | 66.0% | 21.9% | 901.6 | 899.8 | 1,371.8 | 1,312.5 | 26,644 | 62.2% | 21.7% | 257 | 964.6 | 962.6 | 1,681.4 | 1,503.7 |
|  | Clinically vulnerable | 57,742 | 727 | 42,680 | 73.9% | 33.1% | 545.9 | 544.8 | 1,014.5 | 970.6 | 40,380 | 69.9% | 32.9% | 240 | 594.4 | 593.1 | 1,238.2 | 1,107.4 |
|  | Healthy/other | 69,628 | 452 | 58,135 | 83.5% | 45.0% | 369.8 | 369.1 | 543.6 | 520.0 | 55,660 | 79.9% | 45.4% | 220 | 395.3 | 394.4 | 679.1 | 607.3 |
| Flu vaccine | Received seasonal flu vaccine FY2020/21 | 150,300 | 710 | 116,792 | 77.7% | 90.5% | 525.7 | 524.6 | 881.1 | 842.9 | 110,830 | 73.7% | 90.3% | 630 | 568.4 | 567.3 | 1,094.5 | 978.8 |
|  | Did not receive seasonal flu vaccine FY2020/22 | 19,926 | 913 | 12,306 | 61.8% | 9.5% | 723.2 | 721.7 | 877.6 | 839.7 | 11,854 | 59.5% | 9.7% | 87 | 733.9 | 732.4 | 953.3 | 852.5 |
| Recent acute illness | High | 809 | 3,214 | 22 | 2.7% | 0.0% | - | - | 4,545.5 |  | 21 | 2.6% | 0.0% | - | - |  | 9,523.8 |  |
|  | Medium | 521 | 3,647 | 5 | 1.0% | 0.0% | - | - | - |  | <5 | 0.8% |  | - |  |  |  |  |
|  | Low | 168,896 | 713 | 129,071 | 76.4% | 100.0% | 544.7 | 543.5 | 880.1 | 842.1 | 122,659 | 72.6% | 100.0% | 717 | 584.5 | 583.3 | 1,079.4 | 965.3 |
| Region | East Midlands | 8,830 | 657 | 7,085 | 80.2% | 5.5% | 550.5 | 549.3 | 733.9 | 702.2 | 6,771 | 76.7% | 5.5% | 38 | 561.2 | 560.0 | 974.7 | 871.7 |
|  | East of England | 21,805 | 807 | 17,138 | 78.6% | 13.3% | 630.2 | 628.9 | 910.3 | 870.9 | 16,683 | 76.5% | 13.6% | 113 | 677.3 | 675.9 | 1,138.9 | 1,018.5 |
|  | London | 18,923 | 1,575 | 10,539 | 55.7% | 8.2% | 1,176.6 | 1,174.2 | 1,878.7 | 1,797.5 | 9,782 | 51.7% | 8.0% | 120 | 1,226.7 | 1,224.2 | 2,259.3 | 2,020.5 |
|  | North East | 9,300 | 419 | 7,183 | 77.2% | 5.6% | 348.0 | 347.3 | 863.1 | 825.8 | 6,811 | 73.2% | 5.6% | 24 | 352.4 | 351.6 | 1,057.1 | 945.4 |
|  | North West | 22,898 | 751 | 17,913 | 78.2% | 13.9% | 619.7 | 618.4 | 949.0 | 908.0 | 16,742 | 73.1% | 13.6% | 118 | 704.8 | 703.3 | 1,164.7 | 1,041.6 |
|  | South East | 29,011 | 707 | 22,424 | 77.3% | 17.4% | 588.7 | 587.5 | 869.6 | 832.0 | 21,232 | 73.2% | 17.3% | 129 | 607.6 | 606.3 | 1,135.1 | 1,015.1 |
|  | South West | 21,349 | 436 | 16,940 | 79.3% | 13.1% | 318.8 | 318.1 | 543.1 | 519.6 | 16,247 | 76.1% | 13.2% | 58 | 357.0 | 356.2 | 547.8 | 489.9 |
|  | West Midlands | 22,168 | 623 | 17,381 | 78.4% | 13.5% | 402.7 | 401.9 | 817.0 | 781.6 | 16,388 | 73.9% | 13.4% | 79 | 482.1 | 481.1 | 1,037.3 | 927.7 |
|  | Yorkshire and The Humber | 15,882 | 434 | 12,495 | 78.7% | 9.7% | 320.1 | 319.5 | 560.2 | 536.0 | 12,028 | 75.7% | 9.8% | 38 | 315.9 | 315.3 | 681.7 | 609.7 |
|  | Unknown | 60 |  | - | 0.0% | 0.0% |  |  |  |  | - | 0.0% | 0.0% | - |  |  |  |  |
| Total |  | 170,226 | 733 | 129,098 | 75.8% | 100.0% | 544.5 | 543.4 | 880.7 | 842.6 | 122,684 | 72.1% | 100.0% | 717 | 584.4 | 583.2 | 1080.8 | 966.6 |

## Appendix 5 Comparison of vaccinated and unvaccinated controls pre-vaccination programme

The pairwise matching approach has been developed to include information on the socio-demographic characteristics of individuals who have been vaccinated, together with factors that are likely to be associated with individuals’ exposure risk to COVID-19 including the local prevalence of COVID-19 (as captured by the MSOA of residence) and individuals living arrangements (with those living alone being more likely to have interactions with formal and informal carers). The matching approach also accounts for individuals’ susceptibility to developing COVID-19 and the severity of illness experienced, and factors associated with individuals’ behaviours that might predict their chances of being vaccinated.

A logistic regression model was used to test the statistical-significance of the parameters used in the pairwise matching approach for test positivity for COVID-19. Odd ratios for the regression are presented in Table A5-1. These results demonstrate that the majority of parameters used are significantly associated with individuals risk of developing COVID-19 during the monitoring period for the study.

**Table A5-1.** Adjusted odds ratios generated using a logistic regression model to predict test positivity between days 14 and 41 post vaccination event for vaccinated individuals and their pairwise controls. The model is based on data to day 41 post vaccination for 116,539 match pairs. p-values were set to 0.05 significance level, with further significance levels set to 0.01(*), 0.001(**) and <0.001 (***).

| Reference Group | Parameter | Adjusted Odds Ratio | Upper CI 95% | Lower CI 95% | P value | Sign. |
| --- | --- | --- | --- | --- | --- | --- |
|  | Intercept | 0.03 | 0.01 | 0.13 | 0.00 | *** |
| Unvaccinated | Vaccinated | 0.43 | 0.40 | 0.47 | 0.00 | *** |
| Female: 80-82 matched to 78-79 | Female: 82-83 matched to 78-79 | 1.00 | 0.89 | 1.12 | 0.97 |  |
|  | Male: 80-81 matched to 76-77 | 1.01 | 0.90 | 1.13 | 0.88 |  |
|  | Male: 82-83 matched to 78-79 | 0.99 | 0.88 | 1.11 | 0.81 |  |
| Ethnic Group: White | Ethnic Group: Black | 1.58 | 0.81 | 3.07 | 0.18 |  |
|  | Ethnic Group: Other BAME | 1.98 | 1.54 | 2.56 | 0.00 | *** |
|  | Ethnic Group: Pakistani and Bangladeshi | 2.95 | 1.70 | 5.11 | 0.00 | *** |
|  | Ethnic Group: Unknown | 0.66 | 0.36 | 1.20 | 0.17 |  |
| Health Risk: Low/Unknown | Health Risk: Frail and/or clinically extremely vulnerable | 2.37 | 2.14 | 2.62 | 0.00 | *** |
|  | Health Risk: Clinically vulnerable | 1.61 | 1.46 | 1.78 | 0.00 | *** |
| Living Arrangements: Other | Living Arrangements: Living with children | 1.30 | 0.91 | 1.86 | 0.15 |  |
|  | Living Arrangements: Living alone | 0.88 | 0.79 | 0.97 | 0.01 | ** |
| IMD Quintile 3 | IMD Quintile 1 | 1.29 | 1.13 | 1.47 | 0.00 | *** |
|  | IMD Quintile 2 | 1.06 | 0.93 | 1.21 | 0.37 |  |
|  | IMD Quintile 4 | 0.87 | 0.77 | 0.98 | 0.03 | * |
|  | IMD Quintile 5 | 0.85 | 0.75 | 0.95 | 0.01 | ** |
| Did not receive seasonal flu vaccine | Received seasonal flu vaccine | 0.95 | 0.83 | 1.09 | 0.48 |  |
| No/low non-elective bed use | Medium & High non-elective bed use | 0.34 | 0.08 | 1.41 | 0.14 |  |

There may be other factors that are not accounted for by this matching approach that impact the outcomes being monitored. One approach for assessing if such unobserved biases are significant is to monitor outcomes for the intervention and control groups in the period before the intervention to assess how the intervention and control groups comparable. As the outcomes of interest for this evaluation are related to the first case of COVID-19 per person, we cannot do this directly. However, by comparing emergency hospital attendances, emergency hospital stays, outpatient attendances, and planned hospital stays over the period prior to the introduction of the vaccination programme for the vaccinated and control cohorts, we can assess how the matching approach performs in terms of healthcare utilisation (see Figure A5-1).

**Figure A5-1.**
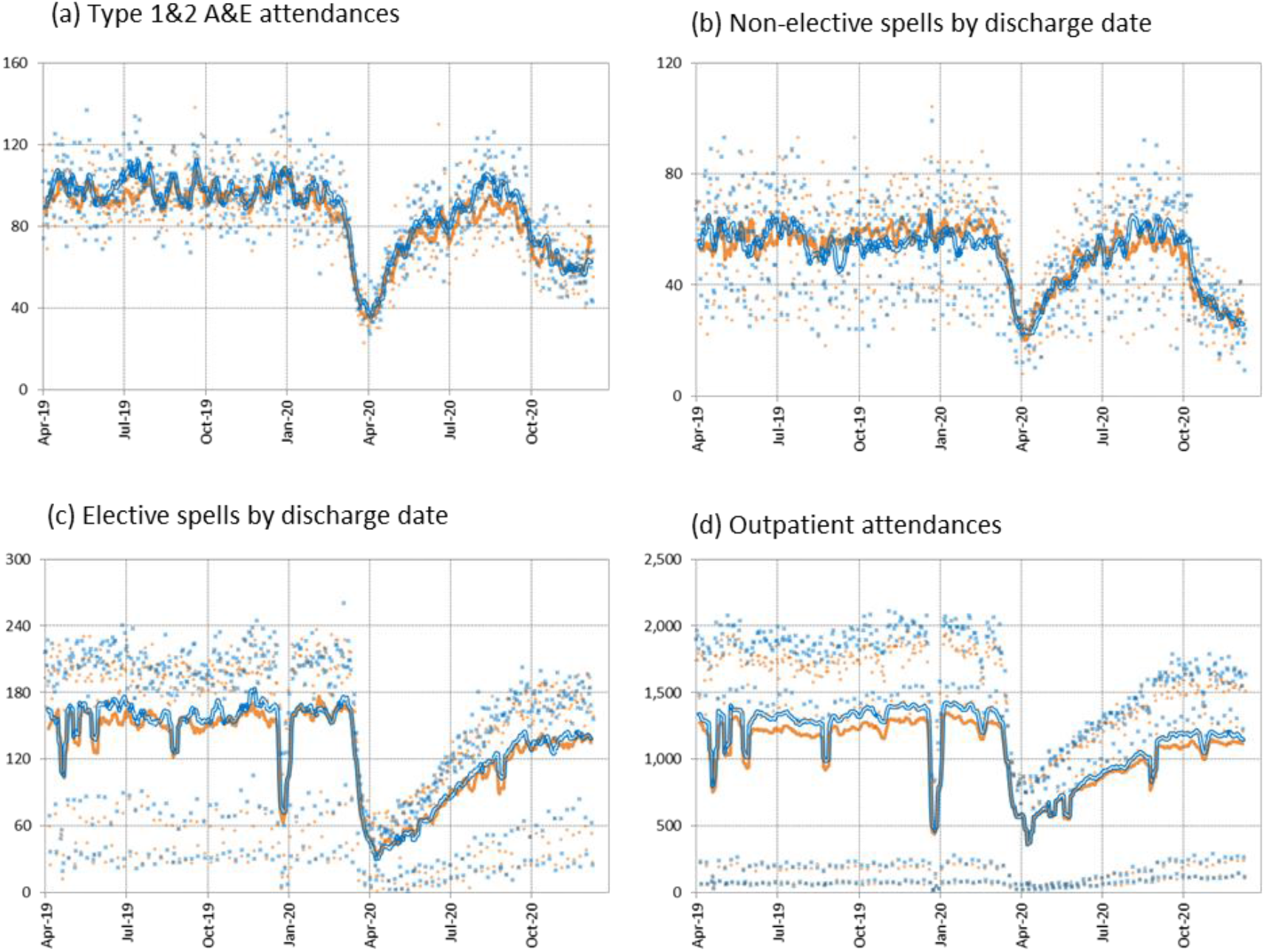
Comparison of the use of hospital-based services per day for the vaccinated group of 80 to 83 years olds (in blue) and the unvaccinated pairwise control group of 76 to 79 years olds (in orange) by activity type. Each data point reflects the daily total, and the solid lines present the seven-day centred moving averages. Data are sourced from the Emergency care Dataset, and the Outpatient and Admitted patient care datasets sourced from the SUS+ service and represent the baseline cohort of 131,236 match pairs at Day 11 post vaccination.

These results demonstrate a close match between the vaccinated and control cohorts in the 18 months to October 2020 for all four activity types. For planned care the vaccinated group is more likely to have used hospital-based health services in the previous 18 months, which is consistent with the vaccinated cohort being an average of 4 years older than their pairwise controls.

The number of negative tests undertaken for individuals within the vaccinated cohort and the control cohort are presented in Figure A5-2. As with the hospital activity measures, the number of negative tests undertaken is similar between the two groups, with the Pillar 1 negative tests (which represents those undertaken following contacts with the health system) showing a closer match when compared to Pillar 2 tests (which represent tests undertaken in the community) where the control group shows higher numbers of negative tests compared to the vaccinated group. This difference in Pillar 2 testing data is consistent with higher uptake rates of Pillar 2 tests by younger age groups, reflecting the average age difference of 4 years between the vaccinated and control groups.

**Figure A5-2.**
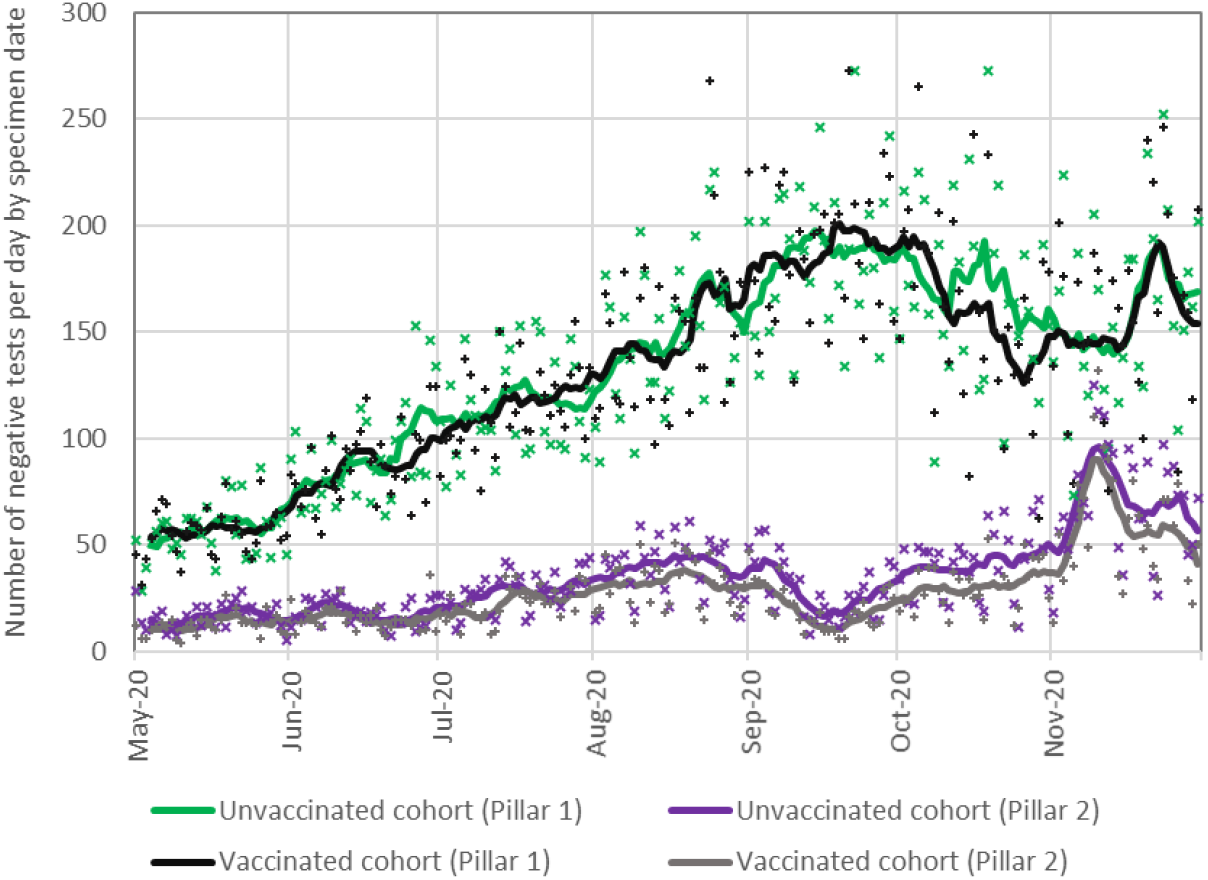
Comparison of the number of negative COVID-19 tests by specimen date undertaken between the 1^st^ May and 30^th^ November 2020 for the vaccinated group and unvaccinated control. Each data point reflects the daily total, and the solid lines present the seven-day centred moving averages.

Overall, Figures A5-1 and A5-2 demonstrate a close match between the vaccinated and control groups before vaccination. These results, couple with the broad range of factors that are accounted for with the pairwise control matching process provides confidence the vaccinated control and matched controls are comparable and can be used to monitor outcomes.

## Appendix 6 Sensitivity of outcomes to control selection

Due to the rapid vaccination rollout, controls who were ineligible for vaccination at the start of the follow-up period relatively quickly become eligible for vaccination (Figure 1 in the main text). This depleted the pool of available controls for matching at later stages of follow-up. In addition, those with documented infection during the study period became concentrated in the control group because, in order to take up vaccination, individuals must not have had documented infection in the preceding two weeks. To adjust for this selection bias, we used a novel adjustment methodology (Methods & Appendix 2).

To test the sensitivity of our results to possible residual selection bias, we repeated analysis matching 80-83-year-old vaccinated individuals to 72-75-year-old unvaccinated controls, as this group became eligible for vaccination later than the control group used in the main analysis (Figure 1). We also tested the sensitivity of the results to using only controls who remained unvaccinated throughout the follow-up period. Finally, for hospital admissions we included an additional outcome measure for patients admitted via A&E based on a disposal code of admitted to hospital, transferred to another provider or died as recorded in the Emergency Care Data Set.

Effectiveness estimates were broadly consistent whichever control population was used (see Figure A6-1 and Table A6-1). Effectiveness estimates were also consistent between the two measures of hospital admission.

**Figure A6-1.**
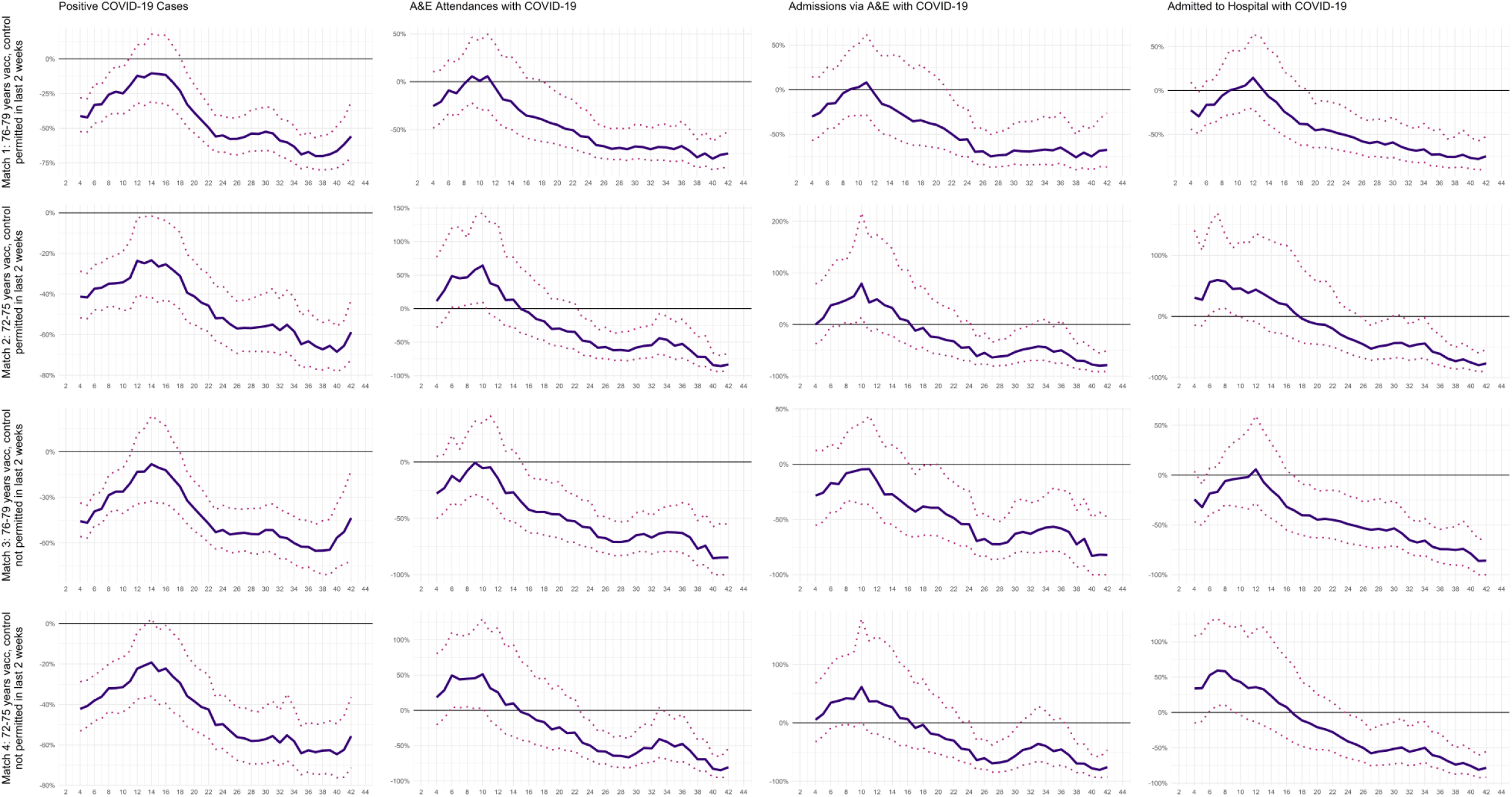
Percentage difference in positive COVID-19 tests, A&E attendances with COVID-19, hospital admission via A&E with COVID-19 and all <43-day length of stay hospital admissions with COVID-19 for four matching strategies by day since first vaccine dose. 95% confidence intervals are represented by the dashed lines.

**Table A6-1.** Comparison of estimates of the effectiveness of the BNT162b2 mRNA Covid-19 vaccine by days since vaccination for four matching strategies.

|  |  | Days since first vaccine dose |  |  |  |
| --- | --- | --- | --- | --- | --- |
| Control/Outcome | Measure | 14 to 20 | 21 to 27 | 28 to 34 | 35 to 41 |
| **Controls aged 76 - 79 years excluding those vaccinated before 14 days of the end of the monitoring period** |  |  |  |  |  |
| Cohort | Size (% Match Rate) | 76.8% | 76.5% | 75.8% | 72.1% |
| Rate of positive COVID-19 tests | Vaccinated group | 28.2 (34.3 to 22.9) | 13.4 (16.2 to 10.7) | 9.7 (11.9 to 7.6) | 4.6 (6.1 to 3.3) |
|  | Unvaccinated group | 34.0 (41.2 to 27.4) | 30.0 (36.7 to 24.1) | 20.9 (26.1 to 16.2) | 15.3 (19.2 to 12.0) |
|  | % Difference | -16.9% (9.4% to -36.7%) | -55.2% (-40.8% to -66.8%) | -53.7% (-35.4% to -66.6%) | -70.1% (-55.1% to -80.1%) |
| Rate of COVID-19 realated A&E attendances | Vaccinated group | 9.0 (12.2 to 6.5) | 5.3 (7.3 to 3.6) | 4.3 (6.0 to 2.9) | 1.5 (2.3 to 0.7) |
|  | Unvaccinated group | 14.3 (20.4 to 9.4) | 12.6 (17.3 to 8.8) | 13.4 (19.3 to 8.4) | 6.9 (10.2 to 4.4) |
|  | % Difference | -36.8% (2.9% to -60.6%) | -57.8% (-30.8% to -74.5%) | -68.1% (-45.2% to -80.9%) | -78.9% (-60.0% to -89.9%) |
| Rate of COVID-19 related hospital admissions via A&E | Vaccinated group | 6.7 (9.6 to 4.6) | 4.4 (6.4 to 2.9) | 3.4 (5.0 to 2.1) | 1.3 (2.2 to 0.6) |
|  | Unvaccinated group | 10.3 (15.8 to 6.0) | 9.9 (14.5 to 5.9) | 10.9 (16.6 to 5.6) | 5.3 (8.1 to 3.2) |
|  | % Difference | -35.4% (23.6% to -62.3%) | -55.7% (-20.8% to -74.6%) | -69.0% (-35.3% to -84.5%) | -75.8% (-50.5% to -89.9%) |
| Rate of COVID-19 hospital admissions | Vaccinated group | 8.5 (11.2 to 5.9) | 6.4 (8.8 to 4.5) | 4.4 (6.12 to 2.9) | 1.5 (2.4 to 0.8) |
|  | Unvaccinated group | 12.1 (16.8 to 8.0) | 13.1 (18.1 to 8.9) | 12.0 (17.9 to 7.8) | 6.1 (8.9 to 3.8) |
|  | % Difference | -30.0% (15.7% to -56.3%) | -51.0% (-19.9% to -69.5%) | -63.7% (-37.1% to -79.2%) | -75.6% (-52.8% to -87.6%) |
| **Controls aged 76 - 79 years excluding those vaccinated before the end of the monitoring period** |  |  |  |  |  |
| Cohort | Size (% Match Rate) | 75.8% | 72.1% | 64.6% | 43.8% |
| Rate of positive COVID-19 tests | Vaccinated group | 28.6 (34.3 to 23.4) | 14.1 (17.3 to 11.1) | 10.3 (13.2 to 7.7) | 4.4 (6.6 to 2.5) |
|  | Unvaccinated group | 34.7 (41.2 to 28.9) | 29.1 (34.9 to 23.8) | 21.3 (25.1 to 17.9) | 12.7 (15.3 to 10.3) |
|  | % Difference | -17.6% (7.1% to -37.2%) | -51.5% (-35.2% to -63.3%) | -51.6% (-34.3% to -66.2%) | -65.1% (-45.4% to -80.8%) |
| Rate of COVID-19 realated A&E attendances | Vaccinated group | 8.7 (11.7 to 6.2) | 5.1 (7.1 to 3.3) | 4.5 (6.4 to 2.9) | 1.3 (2.5 to 0.5) |
|  | Unvaccinated group | 15.6 (20.3 to 11.6) | 12.3 (16.0 to 8.9) | 12.5 (16.0 to 9.3) | 5.8 (7.8 to 4.2) |
|  | % Difference | -44.3% (-15.8% to -63.3%) | -58.2% (-34.5% to -75.2%) | -63.9% (-43.3% to -79.0%) | -77.0% (-51.9% to -92.1%) |
| Rate of COVID-19 related hospital admissions via A&E | Vaccinated group | 6.3 (9.1 to 4.1) | 4.1 (6.2 to 2.5) | 3.6 (5.5 to 2.1) | 1.2 (2.2 to 0.3) |
|  | Unvaccinated group | 11.0 (15.0 to 7.9) | 9.0 (12.6 to 6.4) | 9.3 (12.8 to 6.5) | 4.3 (6.1 to 2.9) |
|  | % Difference | -42.9% (-8.9% to -66.1%) | -54.3% (-23.7% to -73.9%) | -61.2% (-31.3% to -78.9%) | -72.5% (-41.9% to -93.0%) |
| Rate of COVID-19 hospital admissions | Vaccinated group | 8.6 (11.4 to 6.3) | 6.7 (9.4 to 4.6) | 4.8 (6.8 to 3.2) | 1.6 (2.7 to 0.6) |
|  | Unvaccinated group | 13.4 (17.6 to 9.9) | 13.1 (17.6 to 9.6) | 11.6 (14.9 to 8.4) | 6.4 (8.5 to 4.5) |
|  | % Difference | -35.5% (-1.2% to -58.1%) | -49.2% (-21.1% to -68.5%) | -58.4% (-33.1% to -73.9%) | -75.1% (-51.9% to -90.4%) |
| **Controls aged 72 - 75 years excluding those vaccinated before 14 days of the end of the monitoring period** |  |  |  |  |  |
| Cohort | Size (% Match Rate) | 80.7% | 80.6% | 80.5% | 79.6% |
| Rate of positive COVID-19 tests | Vaccinated group | 28.9 (34.8 to 23.9) | 15.0 (18.3 to 11.8) | 9.4 (11.8 to 7.2) | 5.4 (6.9 to 4.0) |
|  | Unvaccinated group | 40.2 (47.4 to 33.4) | 31.0 (37.6 to 25.7) | 20.9 (25.7 to 16.6) | 16.5 (20.9 to 13.1) |
|  | % Difference | -28.1% (-6.5% to -44.7%) | -51.7% (-36.1% to -64.5%) | -55.1% (-37.2% to -68.5%) | -67.3% (-54.0% to -77.4%) |
| Rate of COVID-19 realated A&E attendances | Vaccinated group | 9.2 (12.3 to 6.7) | 5.6 (8.0 to 3.8) | 4.1 (5.8 to 2.8) | 2.0 (3.1 to 1.2) |
|  | Unvaccinated group | 10.9 (15.3 to 7.3) | 11.2 (15.9 to 7.2) | 9.2 (12.8 to 6.2) | 7.3 (11.2 to 4.0) |
|  | % Difference | -15.6% (38.3% to -44.9%) | -49.5% (-16.0% to -70.4%) | -55.6% (-26.4% to -73.9%) | -71.9% (-42.0% to -85.7%) |
| Rate of COVID-19 related hospital admissions via A&E | Vaccinated group | 6.4 (8.8 to 4.4) | 4.3 (6.2 to 2.8) | 3.2 (4.7 to 2.0) | 1.6 (2.6 to 0.9) |
|  | Unvaccinated group | 7.3 (10.8 to 4.2) | 7.7 (11.3 to 4.6) | 6.2 (9.1 to 3.9) | 5.5 (8.3 to 2.9) |
|  | % Difference | -12.0% (70.1% to -47.8%) | -43.9% (8.5% to -69.6%) | -48.5% (-6.5% to -71.9%) | -70.1% (-36.6% to -86.5%) |
| Rate of COVID-19 hospital admissions | Vaccinated group | 9.6 (12.8 to 7.0) | 6.6 (8.7 to 4.6) | 3.9 (5.5 to 2.5) | 1.9 (2.8 to 1.1) |
|  | Unvaccinated group | 9.0 (13.9 to 5.4) | 10.2 (14.6 to 6.3) | 6.9 (11.0 to 4.3) | 7.0 (10.3 to 4.2) |
|  | % Difference | 6.3% (101.5% to -36.9%) | -35.6% (11.4% to -60.2%) | -43.4% (2.0% to -68.1%) | -73.3% (-50.8% to -86.1%) |
| **Controls aged 72 - 75 years excluding those vaccinated before the end of the monitoring period** |  |  |  |  |  |
| Cohort | Size (% Match Rate) | 80.4% | 79.6% | 77.7% | 70.9% |
| Rate of positive COVID-19 tests | Vaccinated group | 28.6 (34.8 to 23.2) | 14.8 (18.0 to 11.8) | 9.2 (11.7 to 7.1) | 5.5 (7.3 to 3.9) |
|  | Unvaccinated group | 38.8 (45.6 to 32.6) | 29.5 (34.9 to 24.9) | 20.7 (25.1 to 16.7) | 14.9 (17.7 to 12.2) |
|  | % Difference | -26.2% (-4.7% to -43.8%) | -49.7% (-34.8% to -61.8%) | -55.6% (-38.5% to -67.5%) | -62.8% (-49.3% to -74.4%) |
| Rate of COVID-19 realated A&E attendances | Vaccinated group | 8.9 (11.7 to 6.4) | 5.2 (7.1 to 3.6) | 3.5 (5.1 to 2.3) | 2.0 (3.0 to 1.1) |
|  | Unvaccinated group | 10.4 (15.7 to 6.6) | 10.4 (15.0 to 6.6) | 7.6 (11.1 to 4.6) | 6.4 (9.5 to 4.1) |
|  | % Difference | -13.9% (44.2% to -49.3%) | -49.9% (-14.7% to -70.3%) | -53.3% (-19.5% to -73.8%) | -69.4% (-41.8% to -85.7%) |
| Rate of COVID-19 related hospital admissions via A&E | Vaccinated group | 6.3 (8.8 to 4.2) | 4.0 (5.8 to 2.6) | 2.9 (4.3 to 1.8) | 1.5 (2.6 to 0.7) |
|  | Unvaccinated group | 6.9 (10.6 to 4.2) | 7.5 (11.0 to 4.6) | 5.4 (8.2 to 3.3) | 4.8 (7.3 to 3.0) |
|  | % Difference | -8.6% (64.6% to -45.7%) | -46.1% (2.9% to -68.4%) | -46.3% (5.0% to -72.4%) | -69.5% (-35.6% to -85.0%) |
| Rate of COVID-19 hospital admissions | Vaccinated group | 9.6 (12.7 to 6.9) | 6.2 (8.5 to 4.3) | 3.7 (5.2 to 2.4) | 1.8 (3.0 to 1.0) |
|  | Unvaccinated group | 9.9 (14.4 to 6.5) | 10.5 (15.0 to 6.9) | 7.4 (11.0 to 4.5) | 6.8 (9.7 to 4.5) |
|  | % Difference | -3.1% (65.0% to -43.4%) | -41.1% (-3.1% to -64.8%) | -49.6% (-7.6% to -71.9%) | -73.7% (-50.0% to -87.3%) |

